# Viewing the US presidential electoral map through the lens of public health

**DOI:** 10.1101/2020.08.05.20169003

**Authors:** Tymor Hamamsy, Michael Danziger, Jonathan Nagler, Richard Bonneau

## Abstract

Health, disease, and mortality vary greatly at the county level, and there are strong geographical trends of disease. Healthcare is a top priority for voters in the United States, and it is important to examine the relationship between voting patterns at the county level and health, disease, and mortality. We perform a comprehensive analysis of the relationship between voting patterns and over 150 different public health and wellbeing variables, comparing counties in all states, including counties in 2016 battleground states, and counties in states that flipped from Democrat to Republican from 2012 to 2016. We also investigate county-level health trends over the last 30+ years and find statistically significant relationships between a number of health measures and the voting patterns of counties in presidential elections. Collectively, this data exhibits a strong pattern: counties that voted Republican in the 2016 election counties are “sicker” than those that voted Democrat.

## Introduction

Healthcare is one of the top priorities for voters.^12^ In some polls, substantial fractions of Democratic voters indicate it is their top priority.^3^ This party-specific preference is reflected in the Democratic primary candidates debates, in which candidates devoted more time to healthcare than any other topic.^4^ While still important to Republican voters, it is subordinate to the topics of terrorism, the economy, social security, immigration, and the military, with an 18 point gap between Democratic and Republican voters on the issue of health care costs and 13 point gap on the issue of Medicare reform.^2^

An important prerequisite to understanding the mechanisms driving this significant difference in topical focus is the systematic examination of demographic differences between Republican and Democratic voters.^5^ A recent study demonstrated the contrasting economic realities between Republican and Democratic voters, showing divergence between their economic fortunes and wealth trajectories from 2008 to 2018.^6^ For example, the median household income in Republican districts declined from $55,000 to $53,000, while that of Democrat districts experienced an increase from $54,000 to $61,000. Additionally, Republican states have experienced relative wage stagnation, with income declining 3.6 percent from 2008 78 to 2018. Given these different economic realities for Republican and Democratic districts, our study suggests that voter concern around the economy will vary by state.

Just as the economy varies greatly by county, so too does health, disease, and mortality. Studies examining county-level trends in mortality rates for the different major causes of death show geographical trends.^9^ Disparities have been demonstrated in life expectancy among US counties over the period from 1980 to 2014.^10^ Part of these geographical differences and county-level inequalities are due to deaths of despair (drug overdose, alcoholic liver disease, and suicide deaths), highlighted as a driving factor in the rising midlife morbidity and mortality among white non-Hispanic Americans.^11,12^ Previous studies have shown strong associations between voting patterns, mortality, health, and disease in the 2016 presidential election. Strong associations have additionally been demonstrated between counties that flipped Republican in 2016 (i.e. those that voted Democrat in 2012) and the rising midlife mortality among white non-Hispanic Americans. This demographic was key for Donald Trump’s victory.^13^ It has been shown that Trump over-performed in counties with high drug, alcohol, and suicide mortality rates.^14^ A strong association has been shown between life expectancy and both the proportion of votes in a county that went Republican in 2016 as well as the Republican margin shift from 2012 to 2016. This highlights the diverging life expectancies of Republican and Democratic counties and the possible impact of life expectancy on voting behavior.^15^ Additional associations between several county community health variables and the net Republican voting shift in 2016 have been previously demonstrated.^16^

Investigating the relationship between voting patterns at the county level and health, disease, and mortality in the US is important for framing future narratives surrounding healthcare reform in the context of the 2016 presidential election results and in the context of ongoing 2020 election. While previous studies have looked at the relationship between voting patterns and life expectancy, mortality risk, and public health variables individually, we aim to do a comprehensive analysis of the relationship between voting patterns and over 150 different public health and wellbeing variables. In our analysis, we compared counties in all states, as well as counties in battleground states, and counties in states that flipped from Republican to Democrat from 2012 to 2016, investigating both the relationship of health and wellbeing with the voting margin shifts from 2012 to 2016 as well as overall voting proportions. We believe that investigating associations with shifts in voting and focusing on the battleground and flipped states can provide discontinuities that allow higher-resolution exploration of associations between political and health outcomes. In addition to comparing recent values of these variables with county-level voting patterns, we look at the dynamics of different public health and wellbeing variables over the last 30+ years. We believe that examining these changes over time can both shed light on a changing electorate and elucidate healthcare trends in counties. Our objective is to present data showing the relationship between voting patterns in the US and public health, healthcare, life expectancy and mortality rates at the county level.

## Materials and Methods

In order to show the relationship between over 150 different public health, mortality, and life expectancy variables, with voting patterns at the county level, variables from a number of different publicly available data sources were aggregated and aligned at the county level. The health and wellbeing data as well as their sources include: diabetes, physical inactivity, and obesity crude rates from the Centers for Disease Control and Prevention diabetes surveillance atlas^17^; mortality rate data from the Global Burden of Disease, including county level mortality rates for a number of respiratory diseases, infectious diseases, cardiovascular diseases, cancers, deaths of despair, and mortality risk at different ages as well as life expectancy;^18^ healthcare cost data from the Centers for Medicare and Medicaid Services, including variables such as costs per capita and costs broken down by imaging/drugs/hospice/procedures/dialysis;^19^ Medicaid-relevant data collected from the American Community Survey (ACS), including variables about Medicaid usage at the county level;^20^ disability related data also collected from the ACS; Insurance/Uninsurance rate information collected from the Small Area Health Insurance Estimates (SAHIE);^21^ and a number of public health and demographic variables were collected from the County Health Rankings resource, including health behavioral information (i.e. smoking, drinking, and food indices), access to healthcare, and demographic information, among other data.^22^ All of these variables are presented in Tables 1 and 2 and categorized into the following groups: social, physical and economic environment; respiratory diseases; life expectancy and mortality; insurance and healthcare cost; infectious diseases; health outcomes; health behaviors; demographic; deaths of despair; clinical care; cardiovascular diseases; and cancers.

**Table 1.**
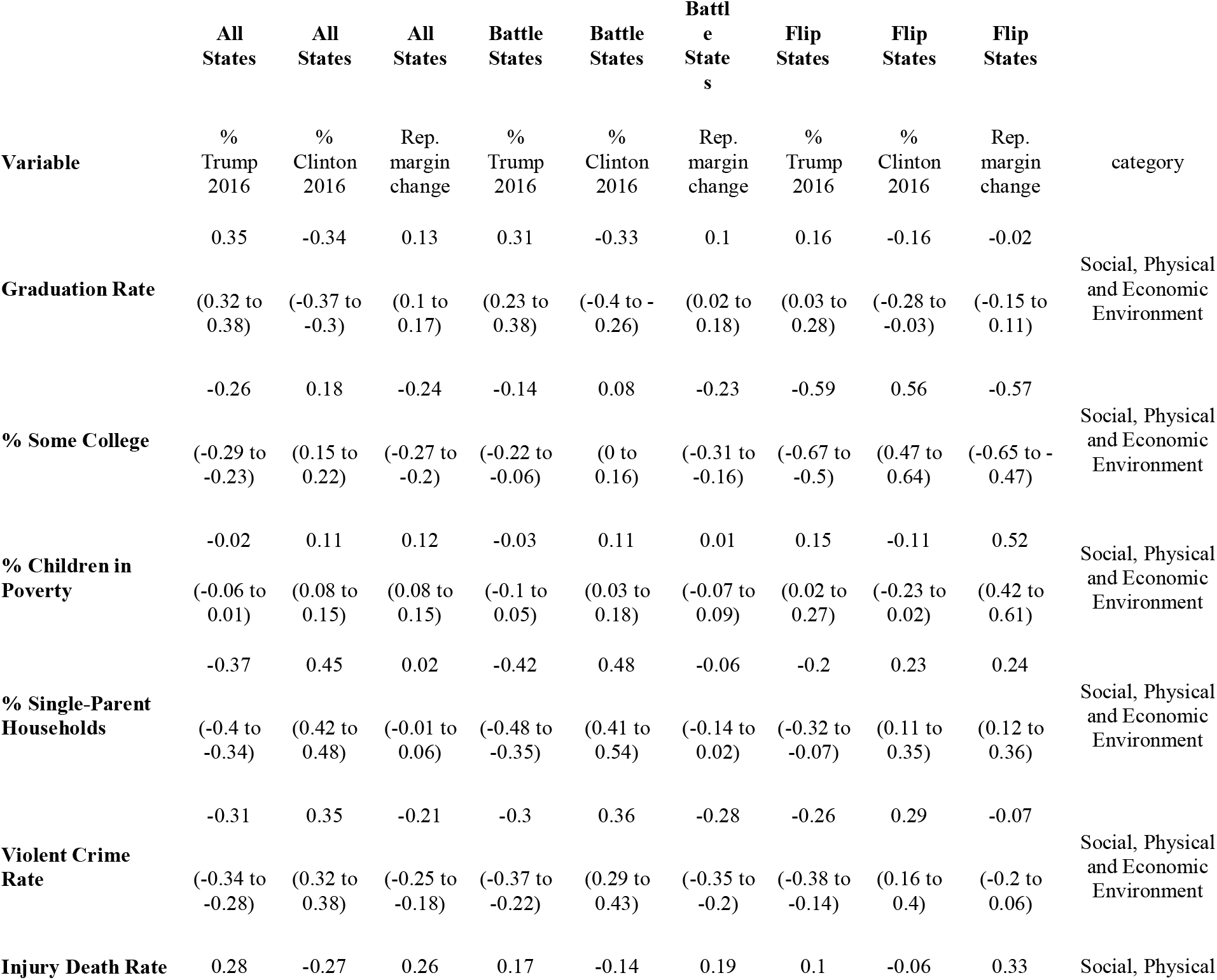
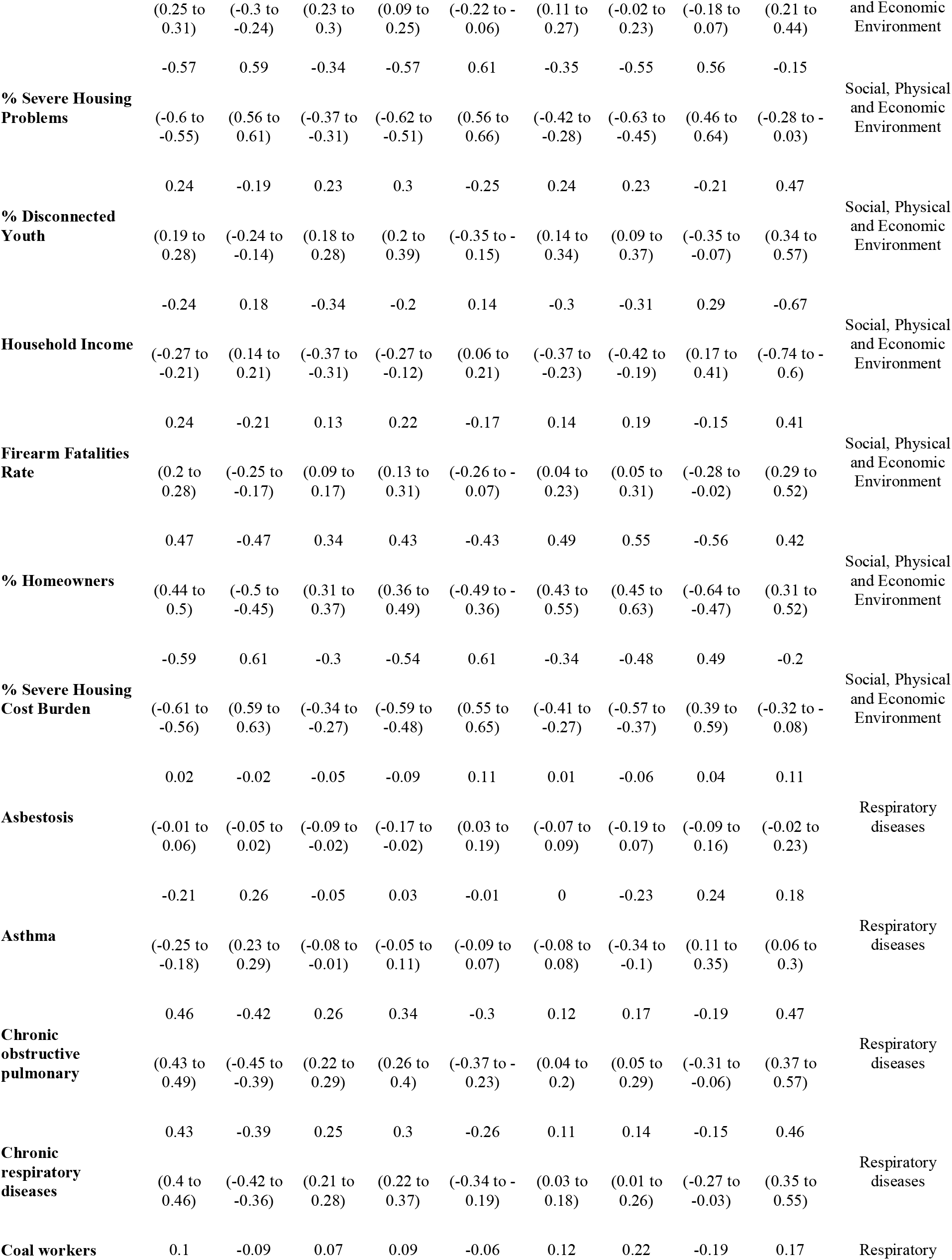
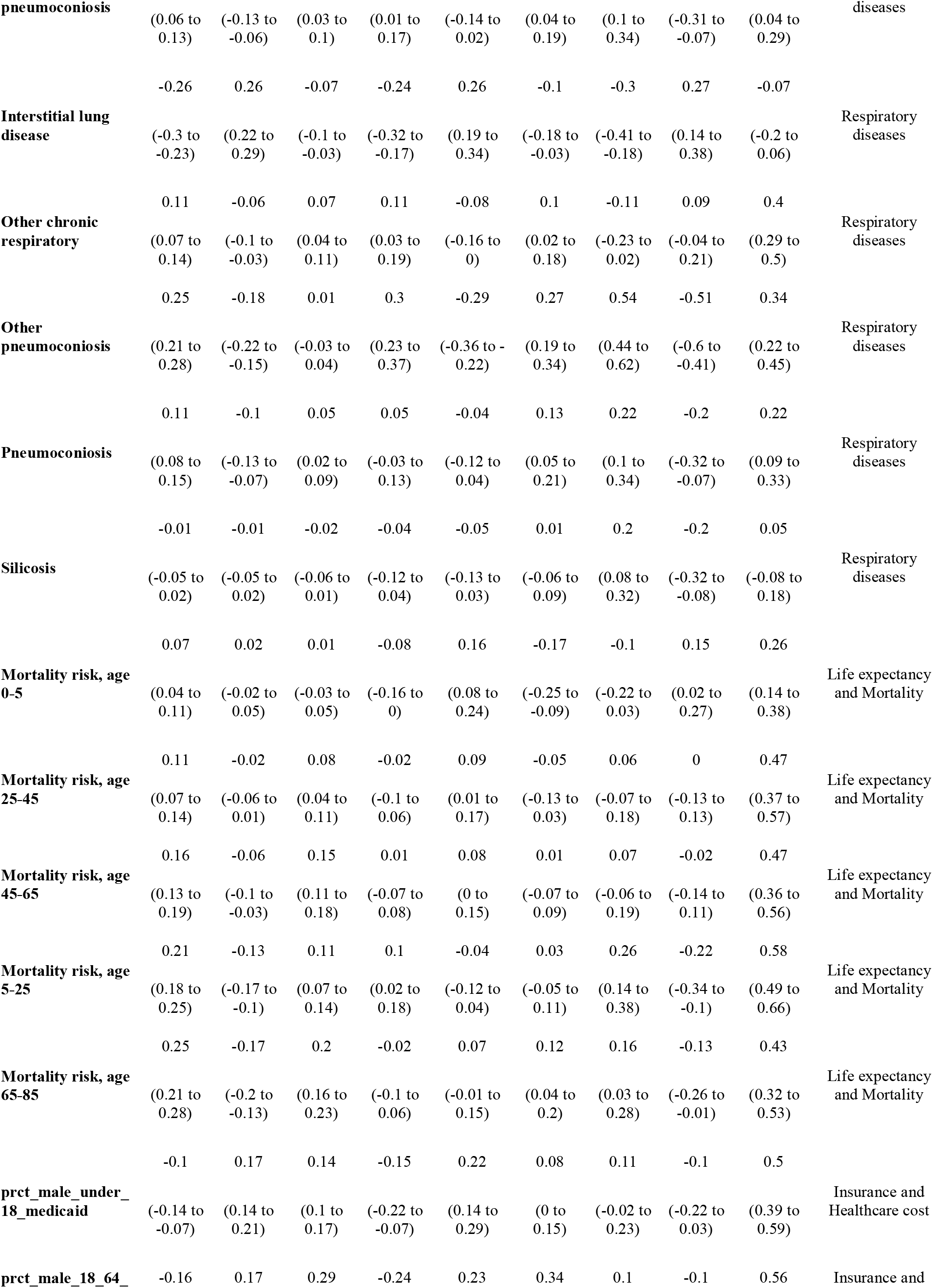
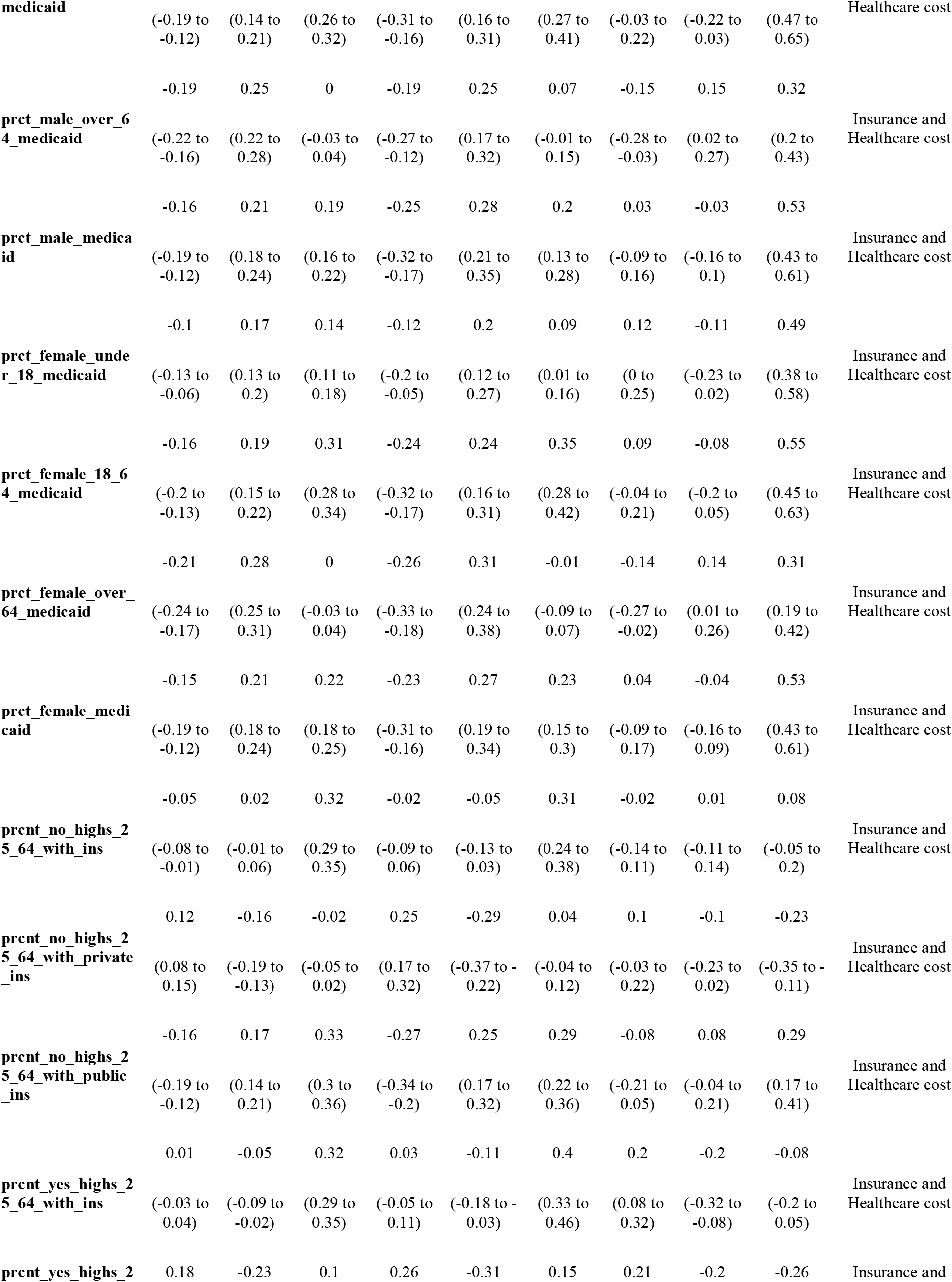
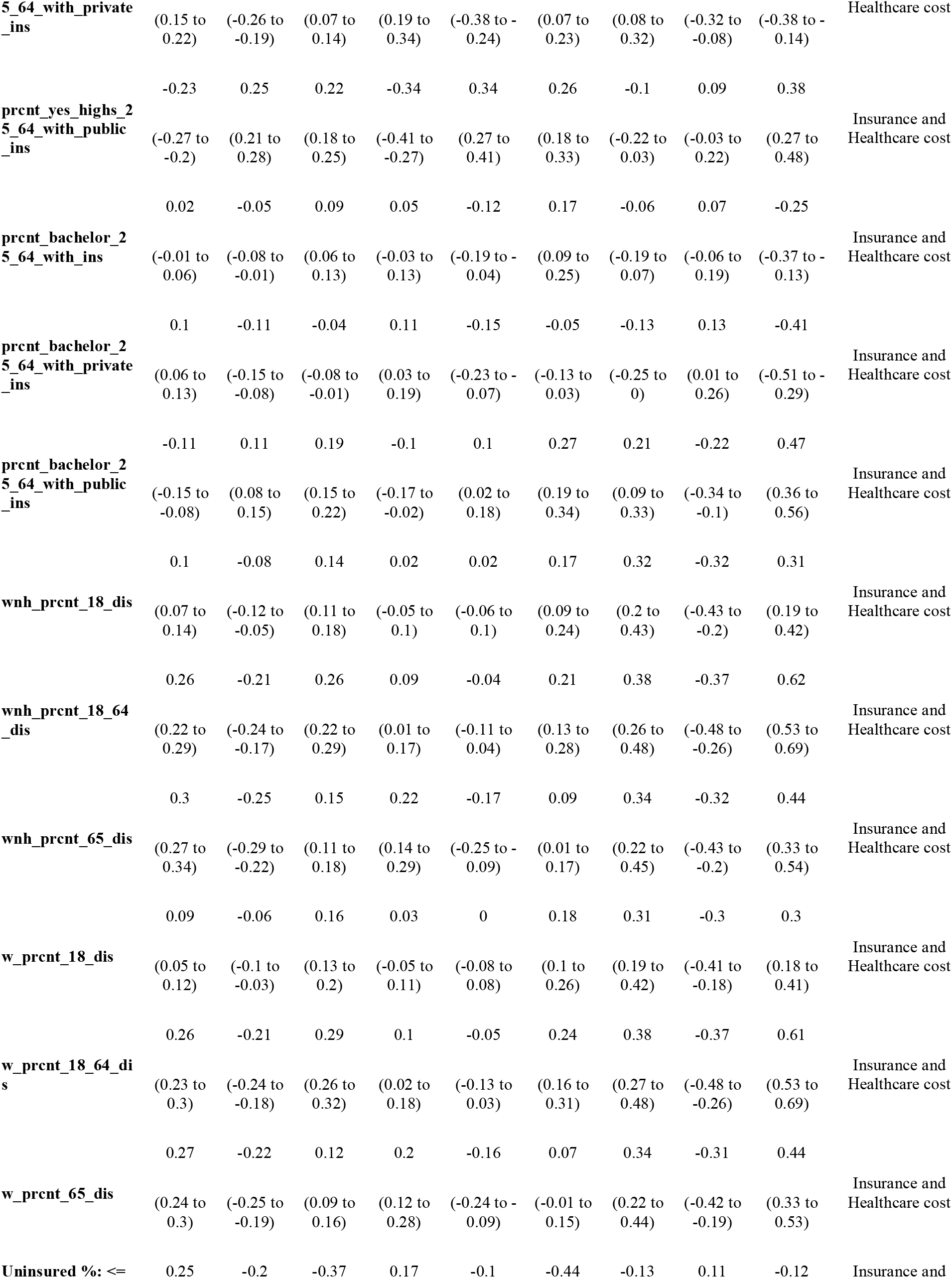
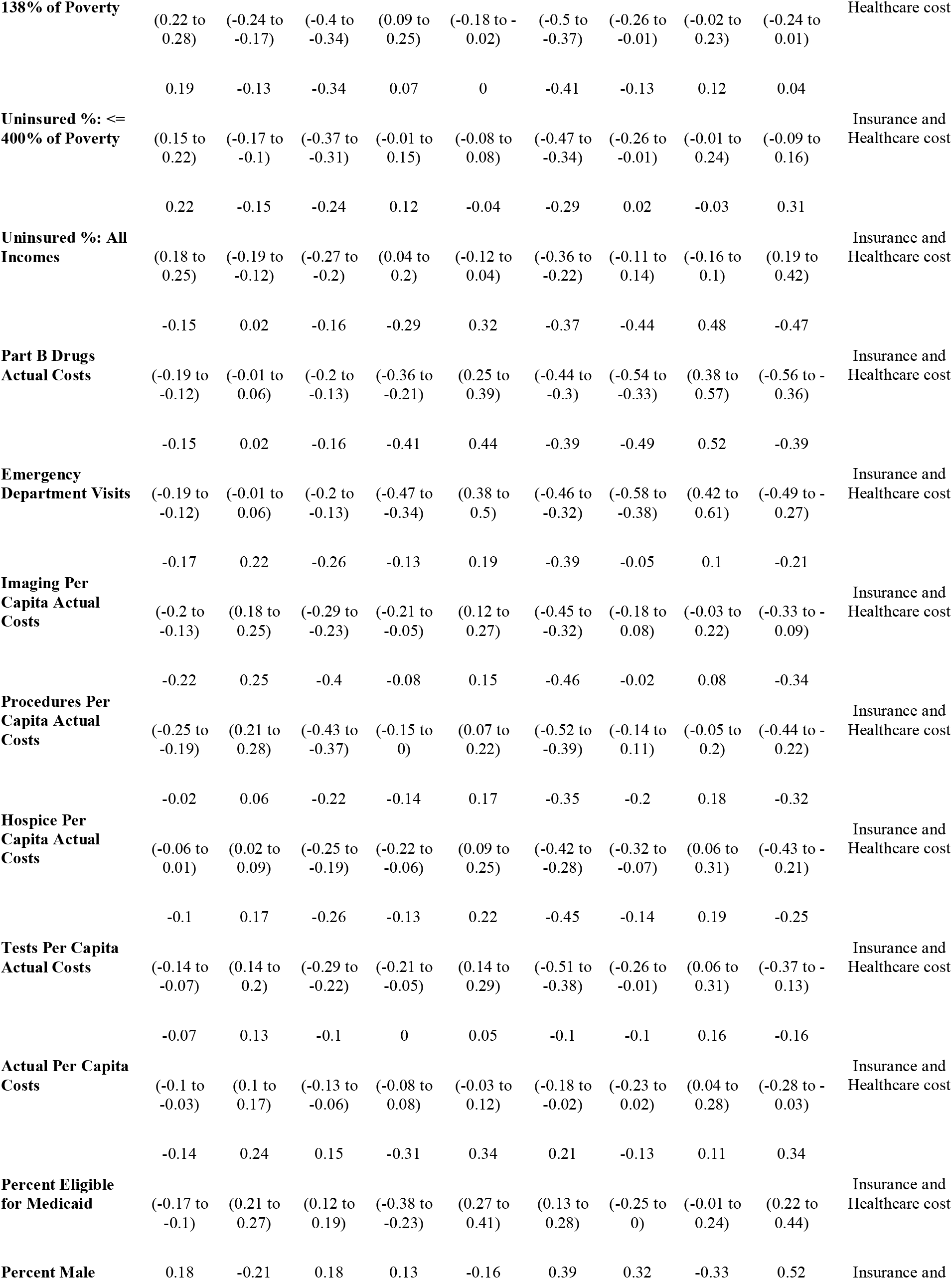
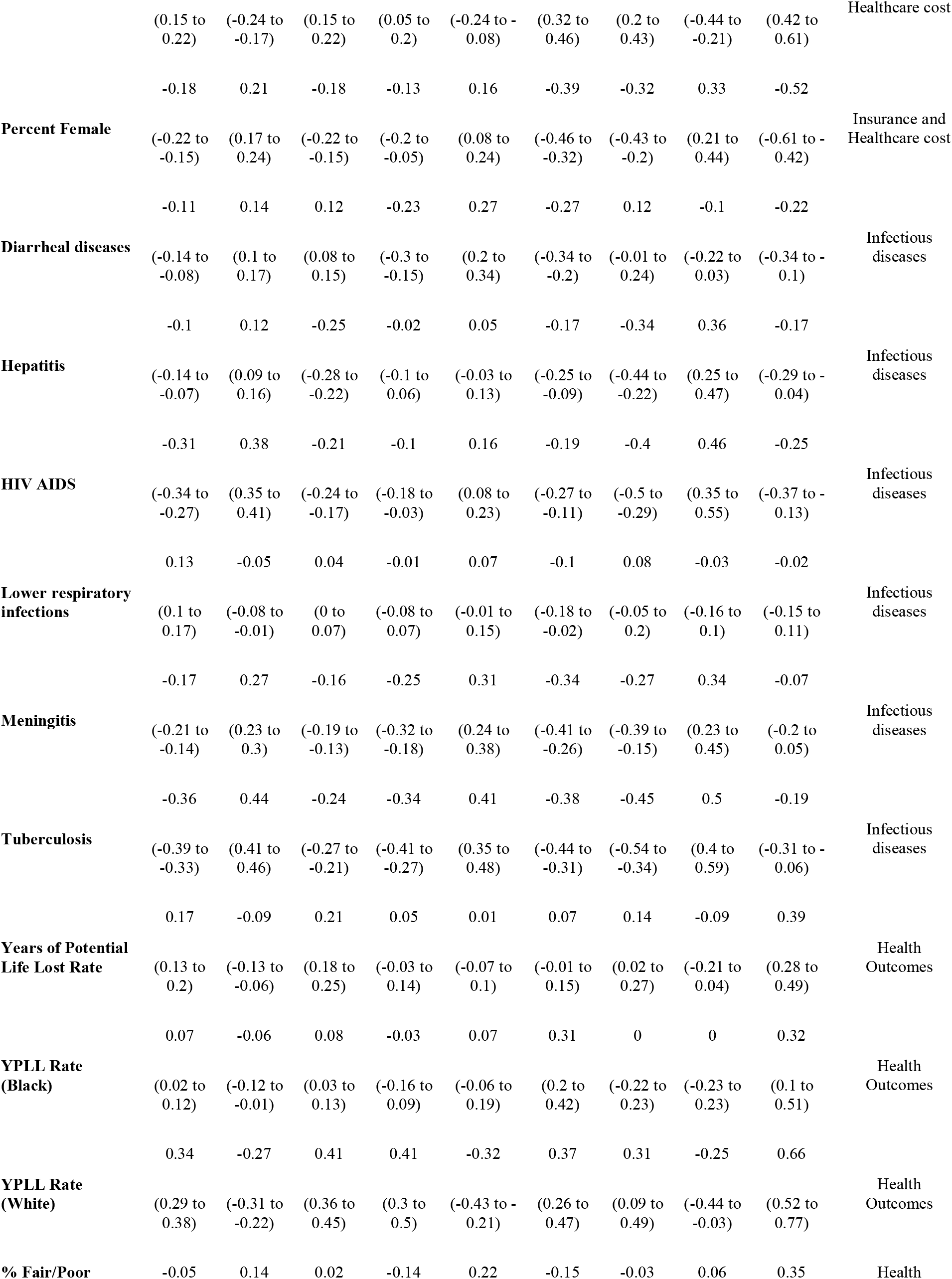
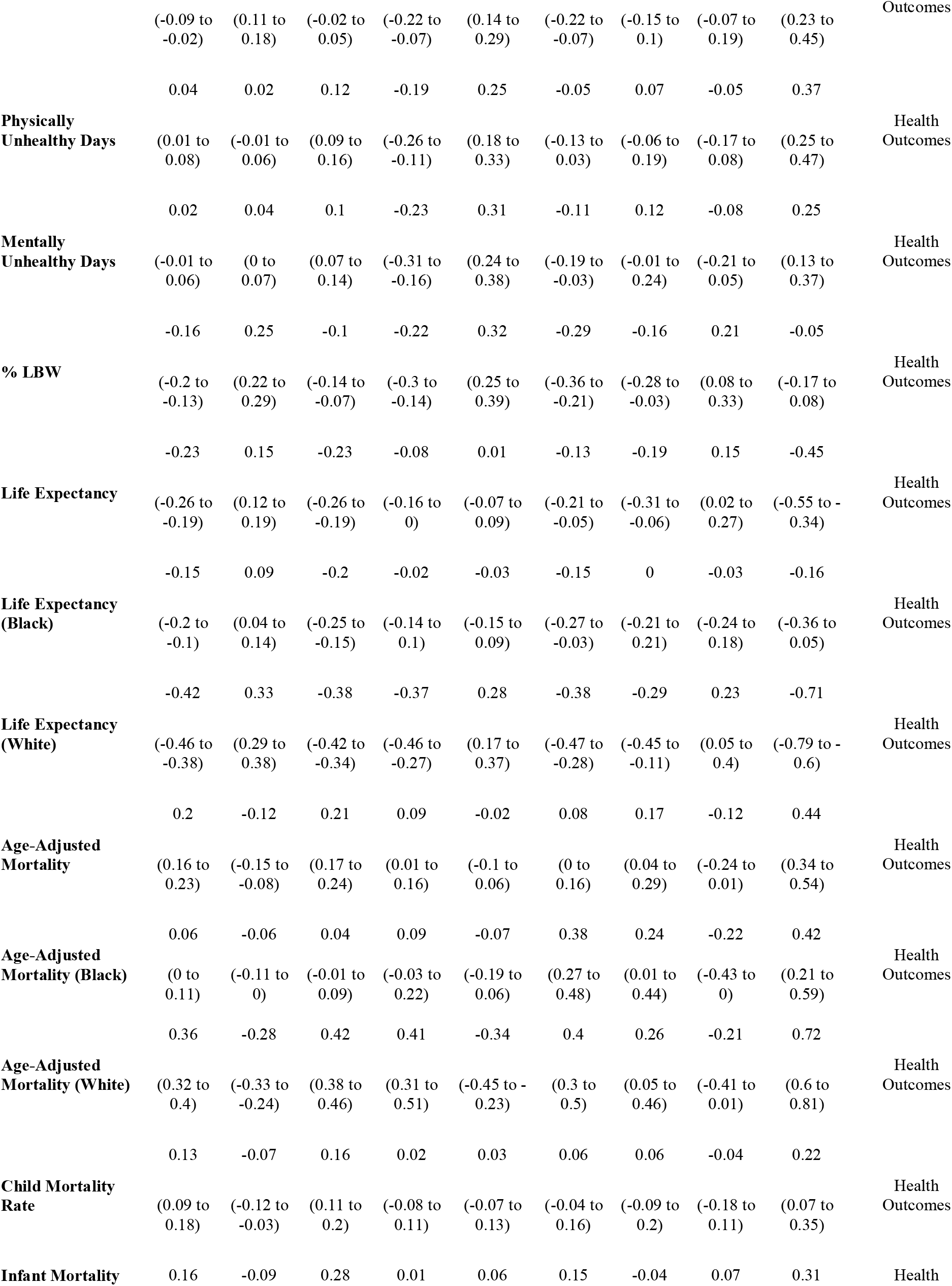
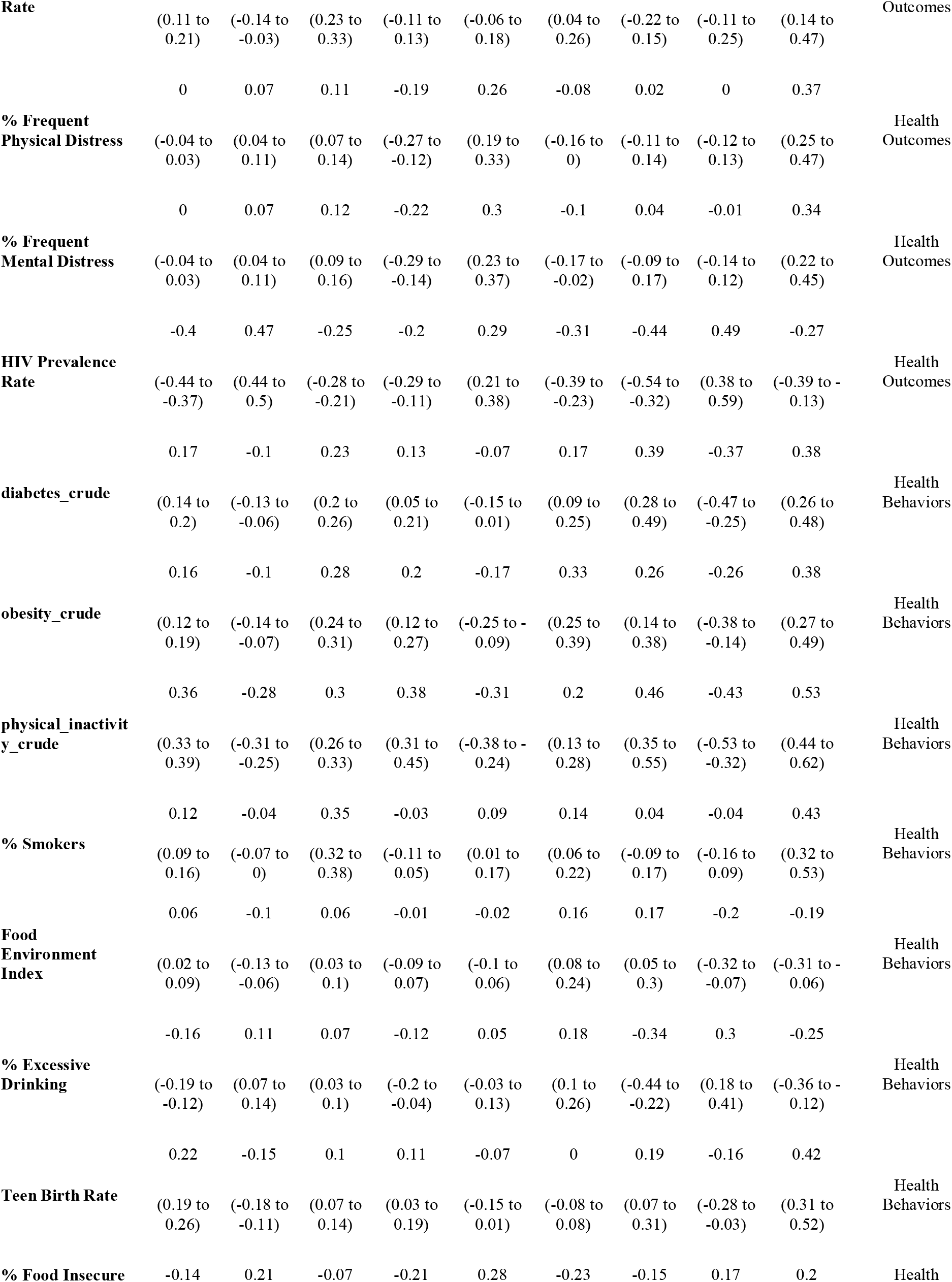
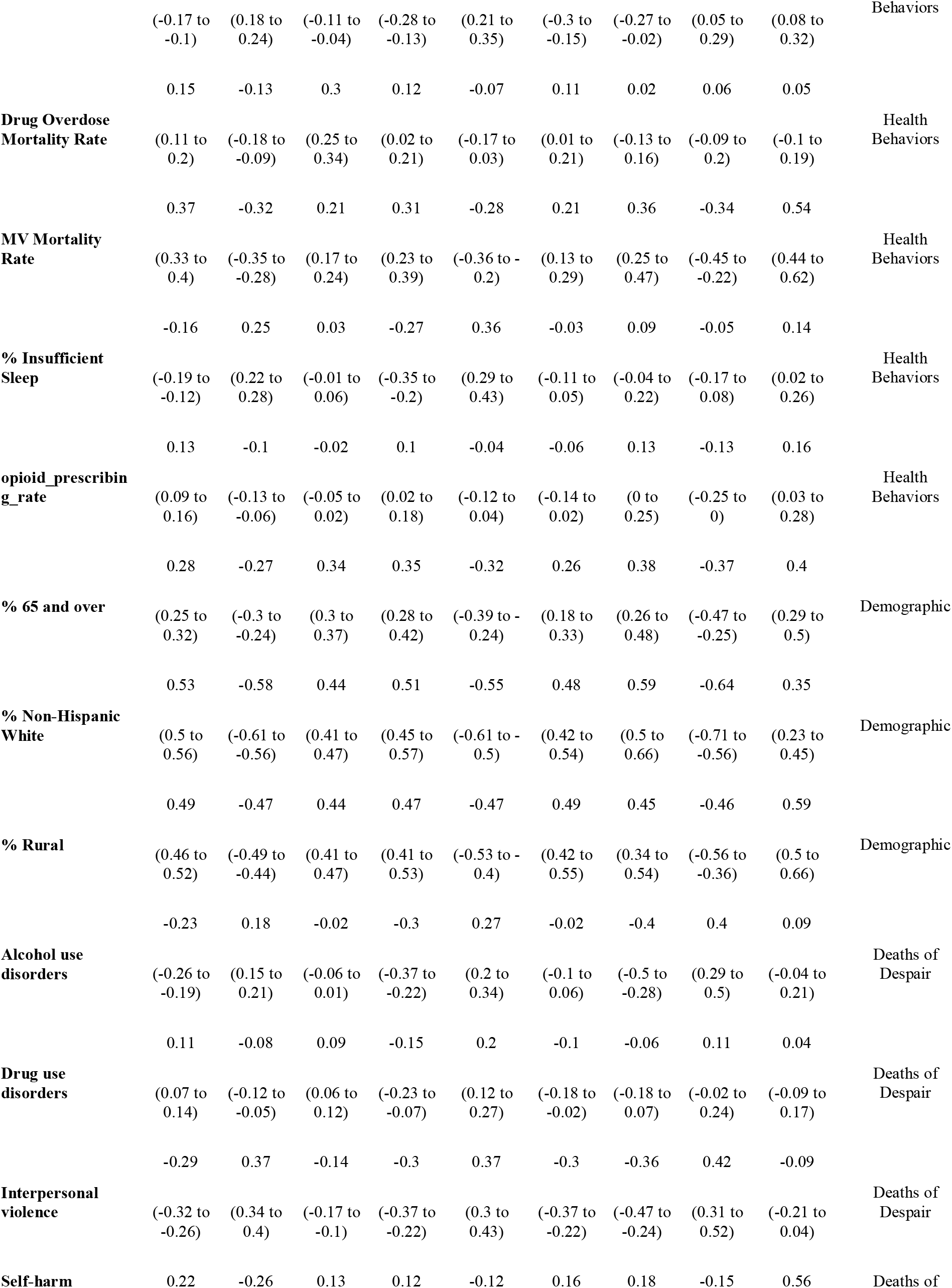
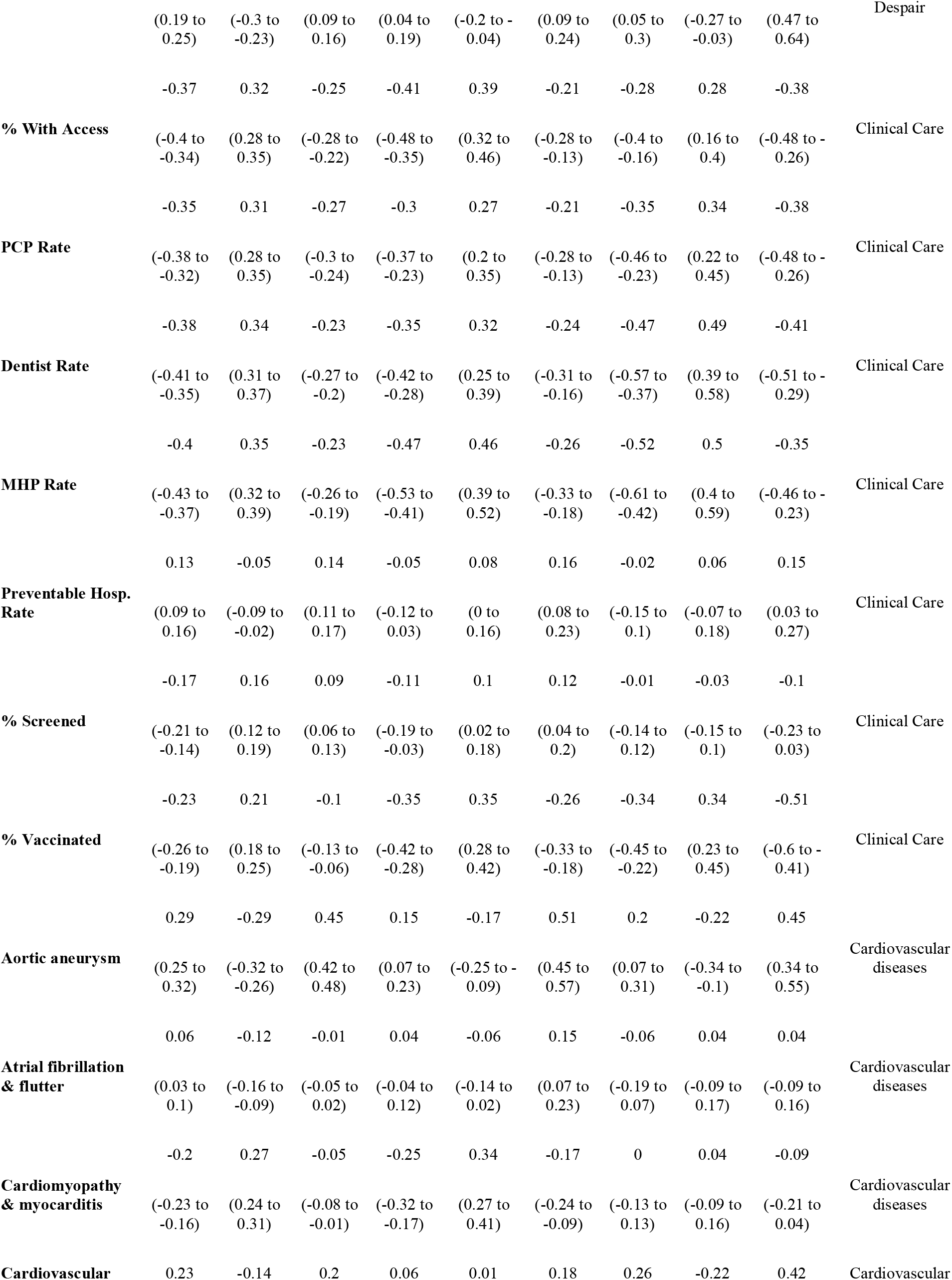
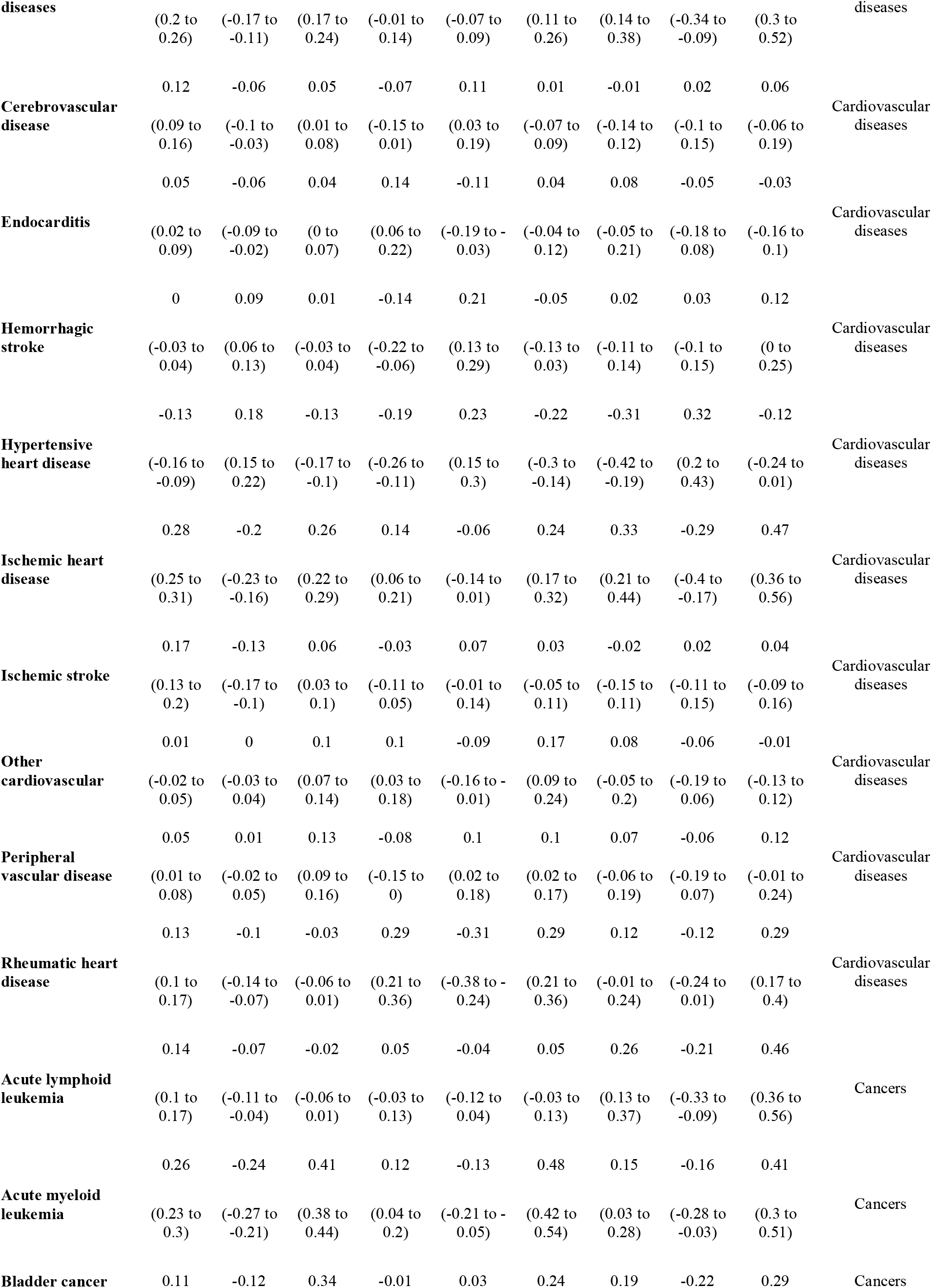
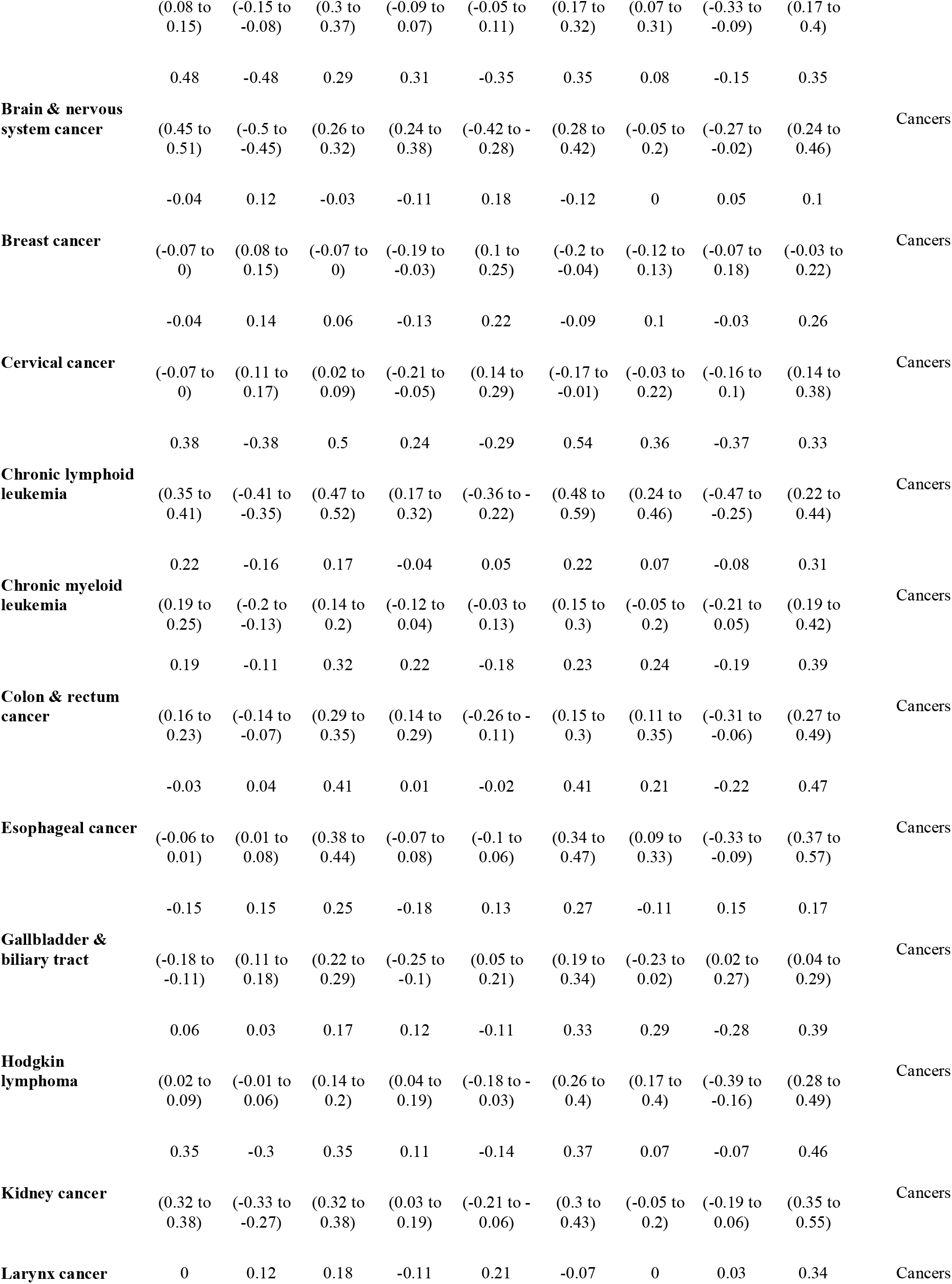
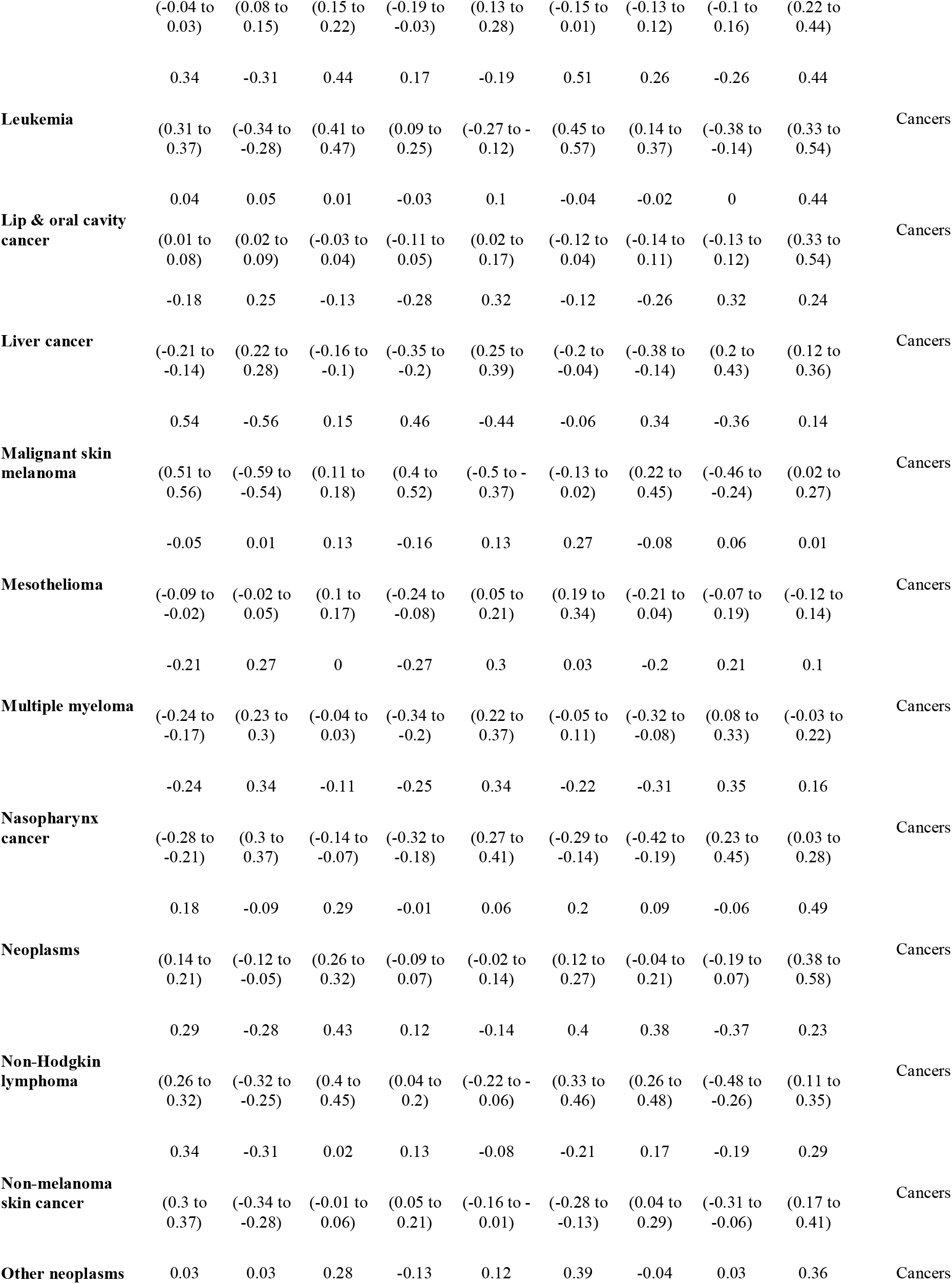
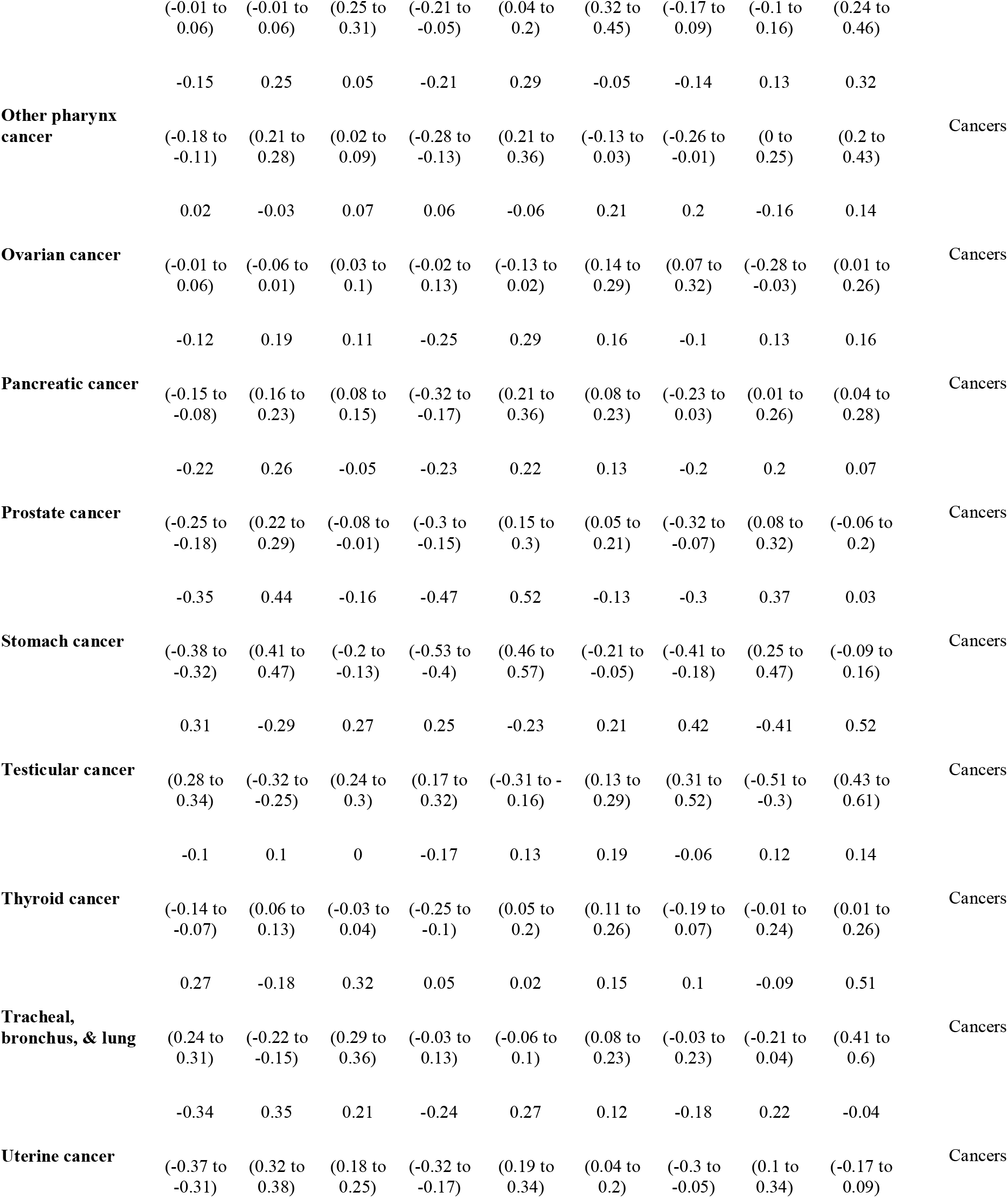
Pearson correlations between different public health-related variables with the percentage of voters in the county that voted for Donald Trump or Hilary Clinton, and the Republican margin shift (from 2012 to 2016). Correlations for counties from all states, counties from battleground states, and counties from states that flipped from Democrat in 2012 to Republican in 2016, are presented.

**Table 2.**
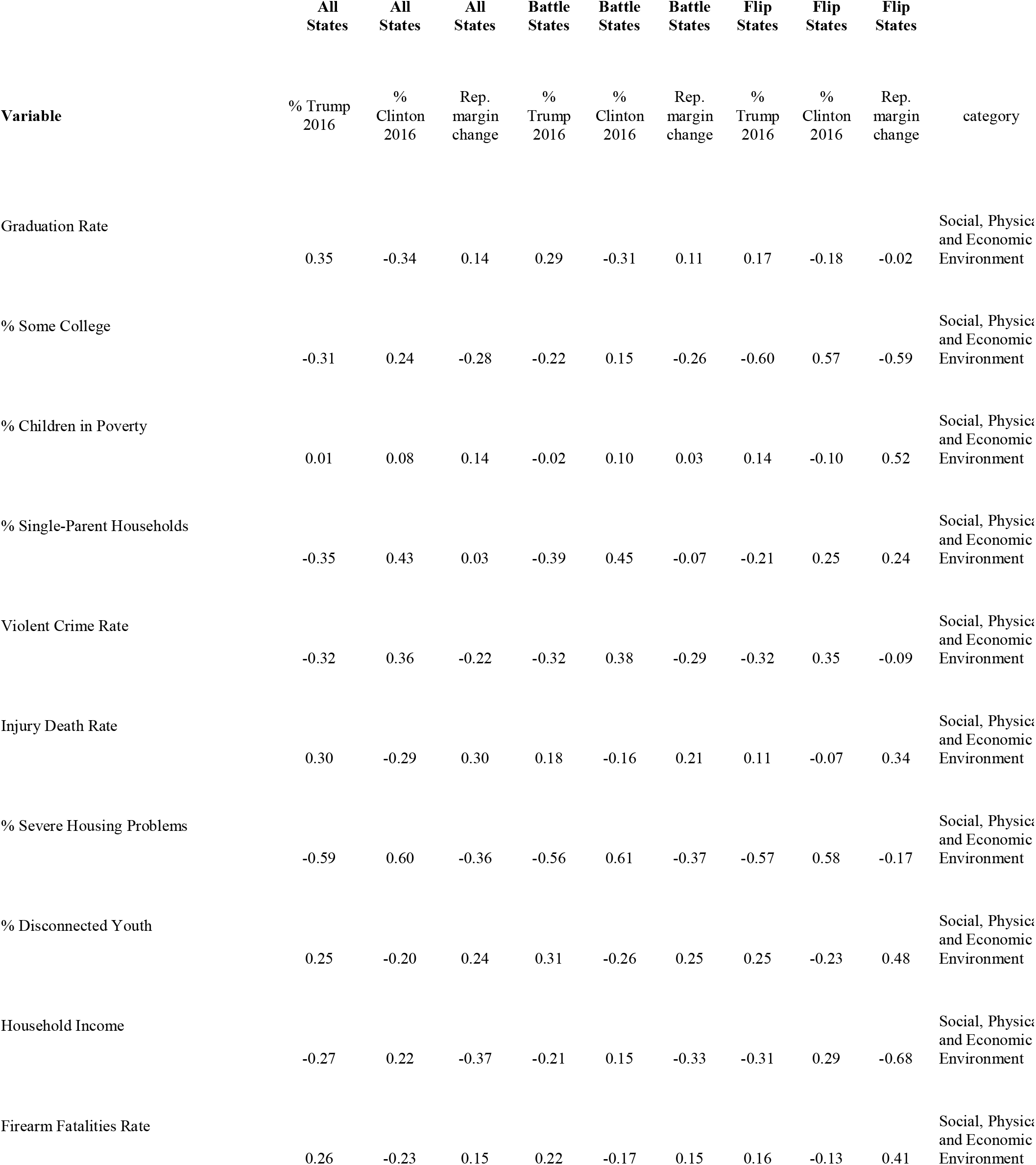
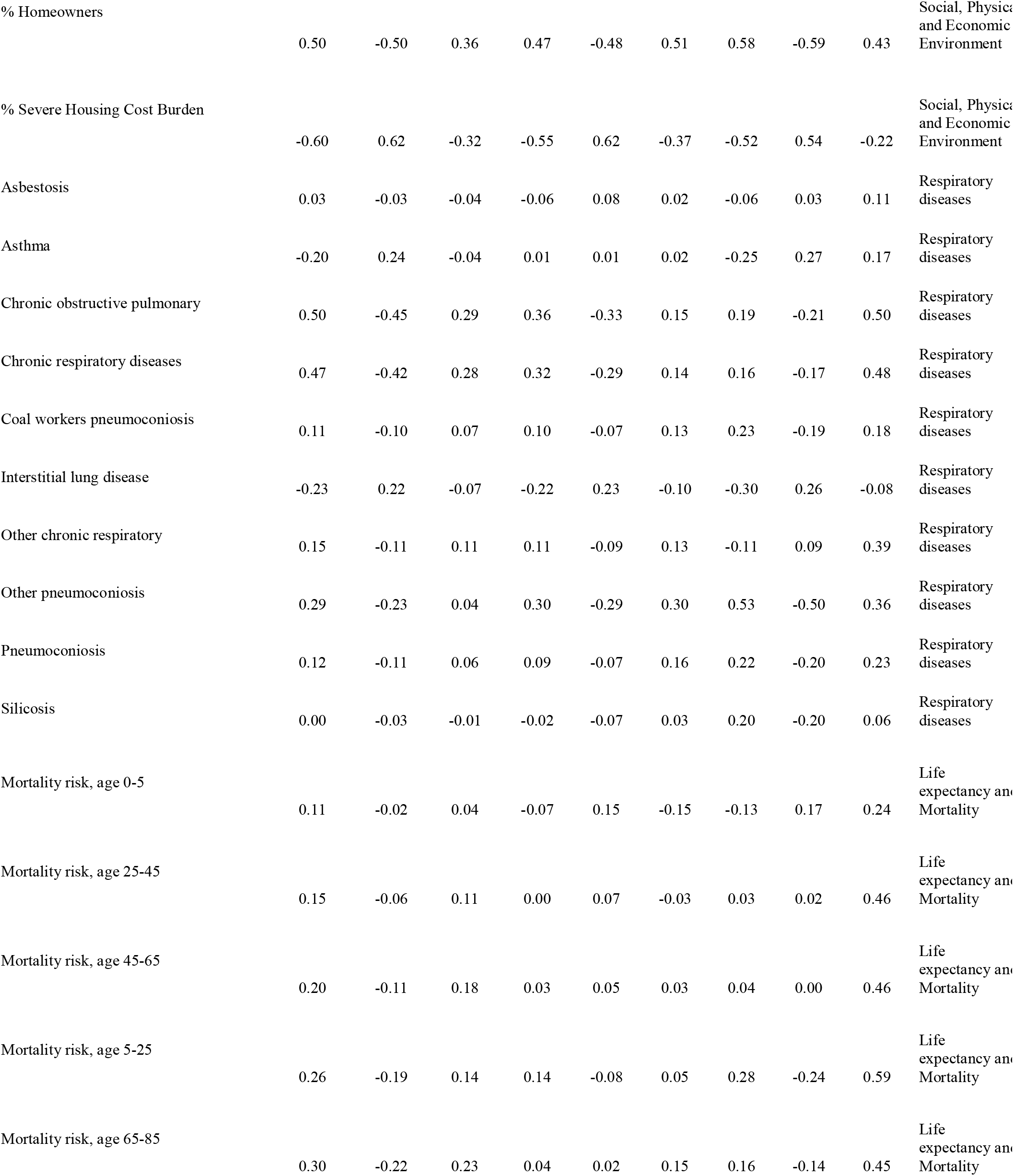
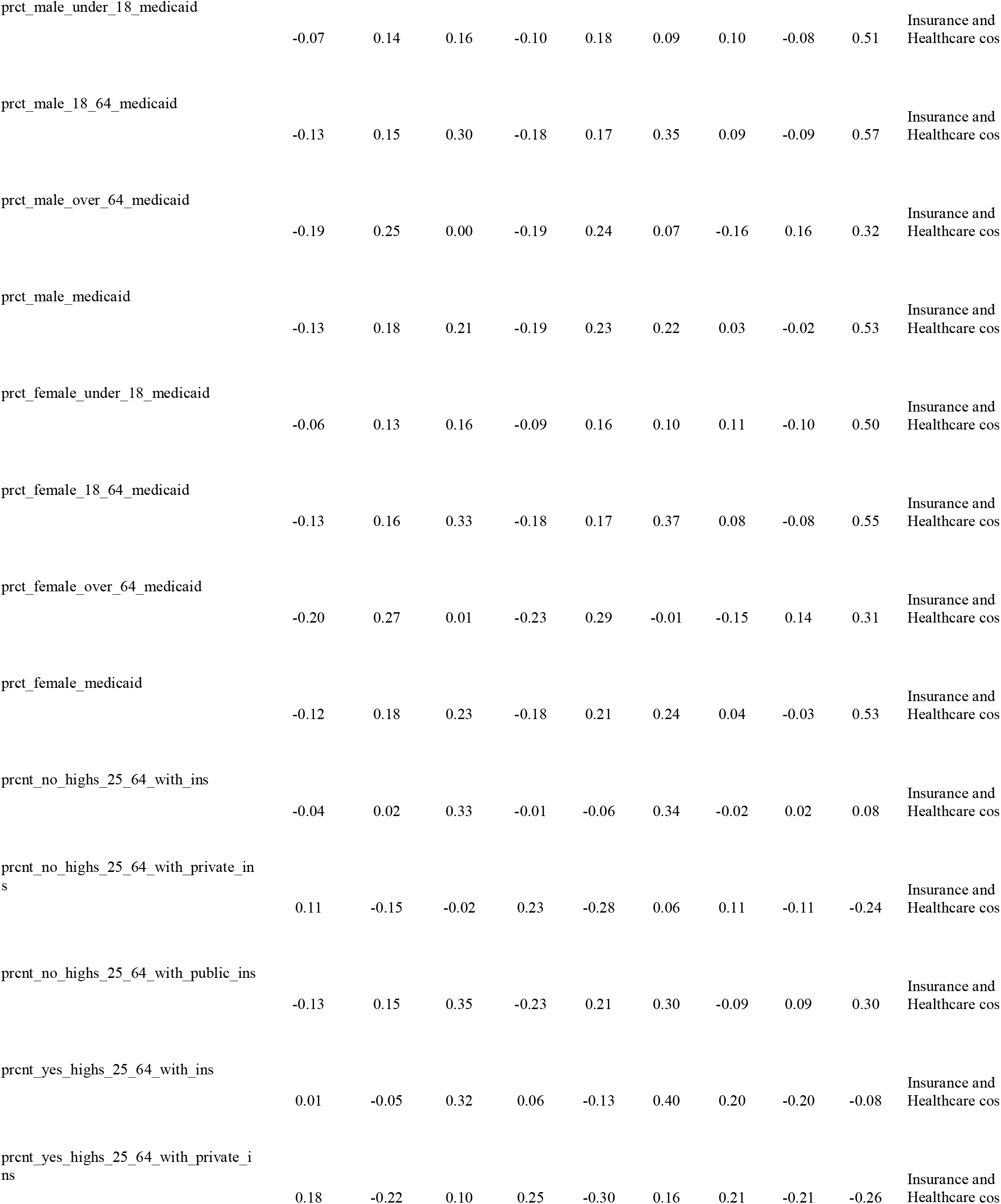
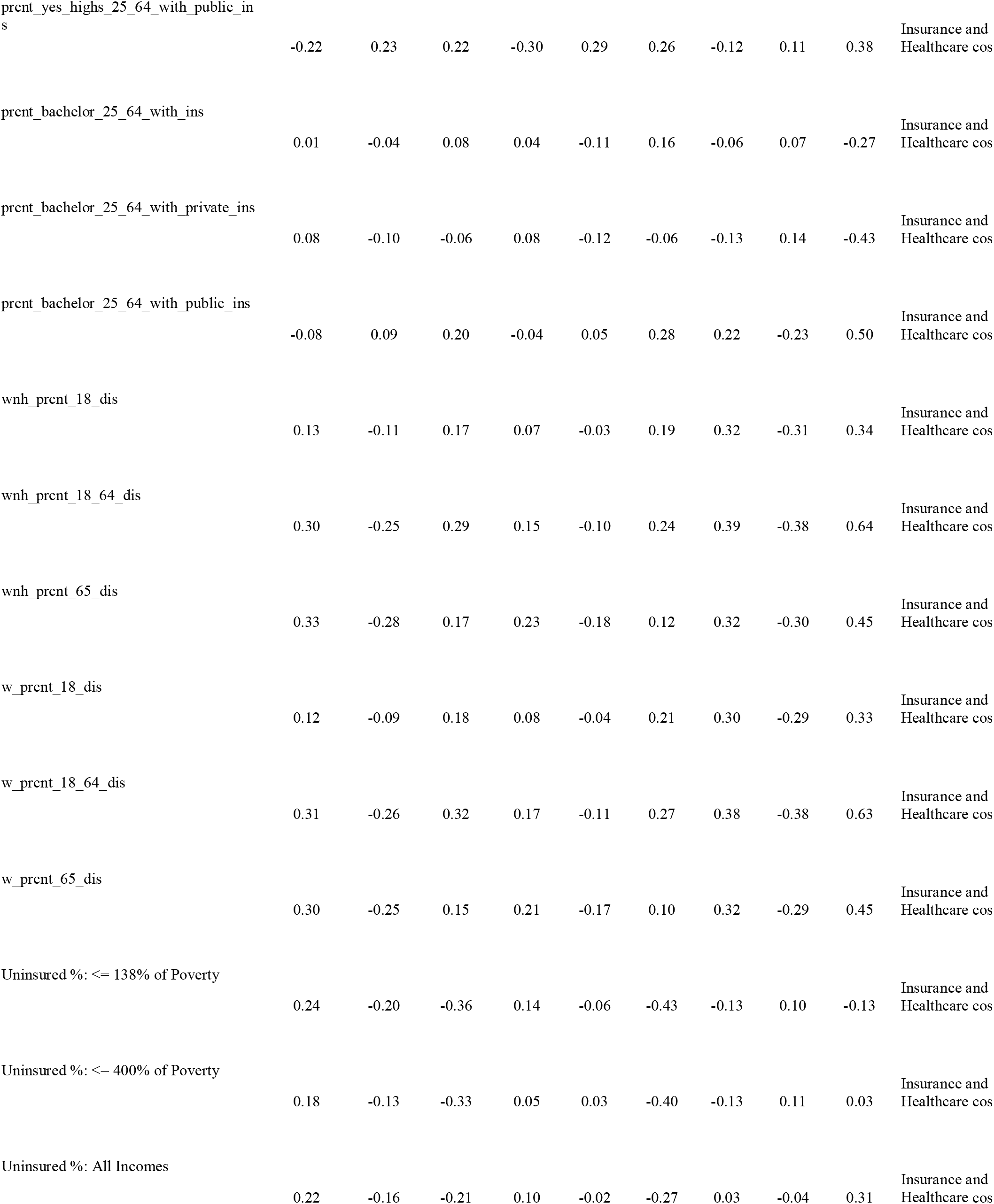
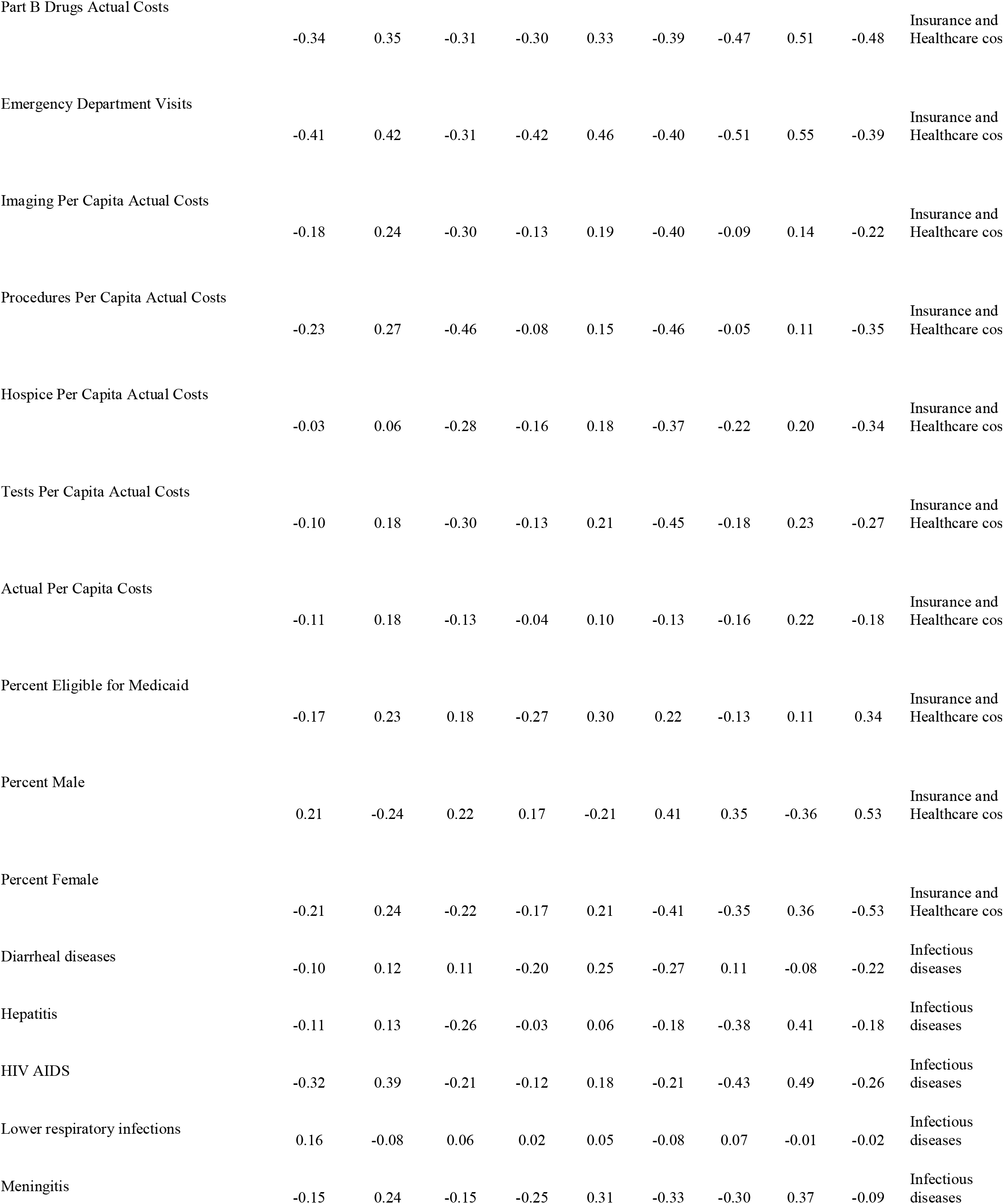
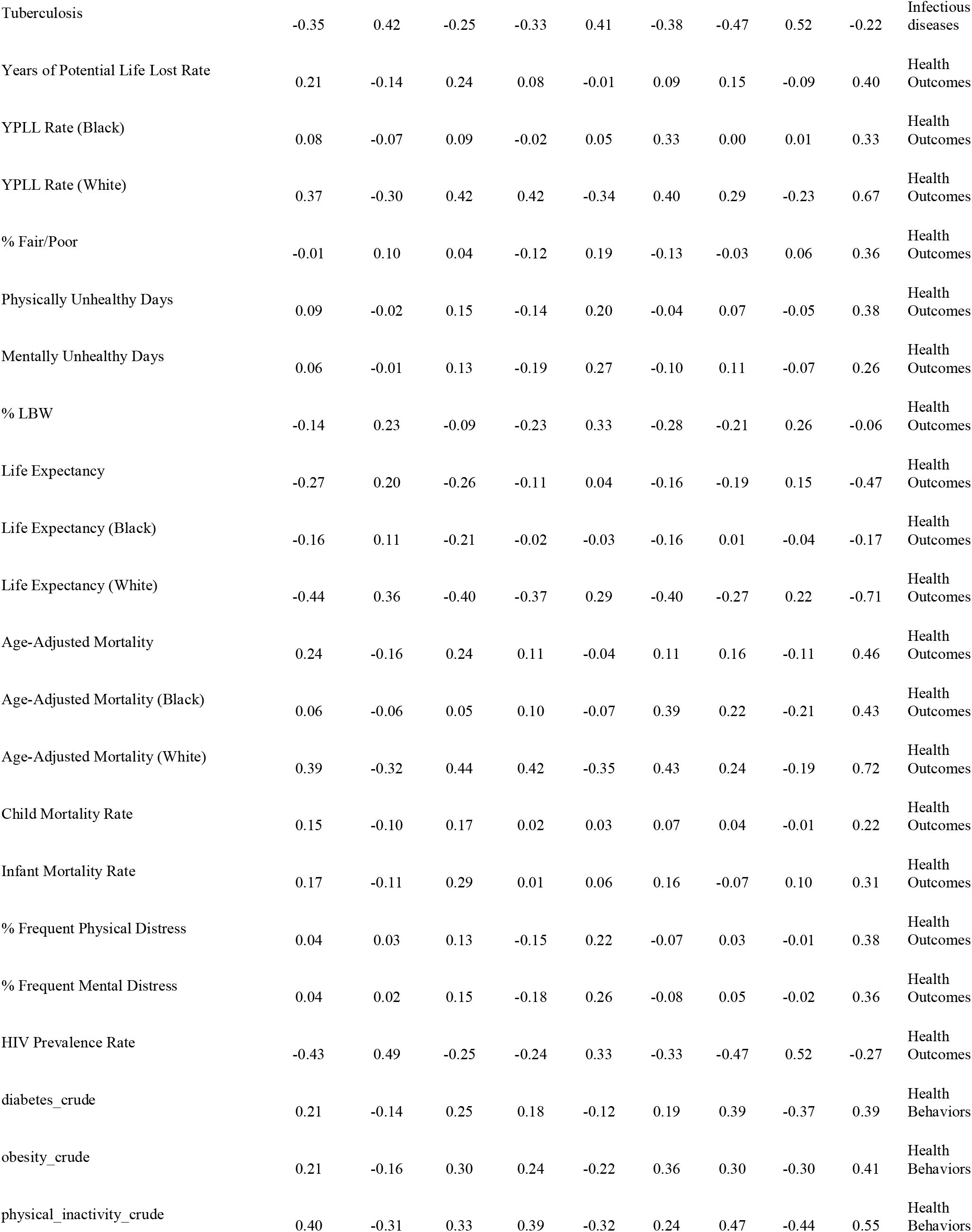
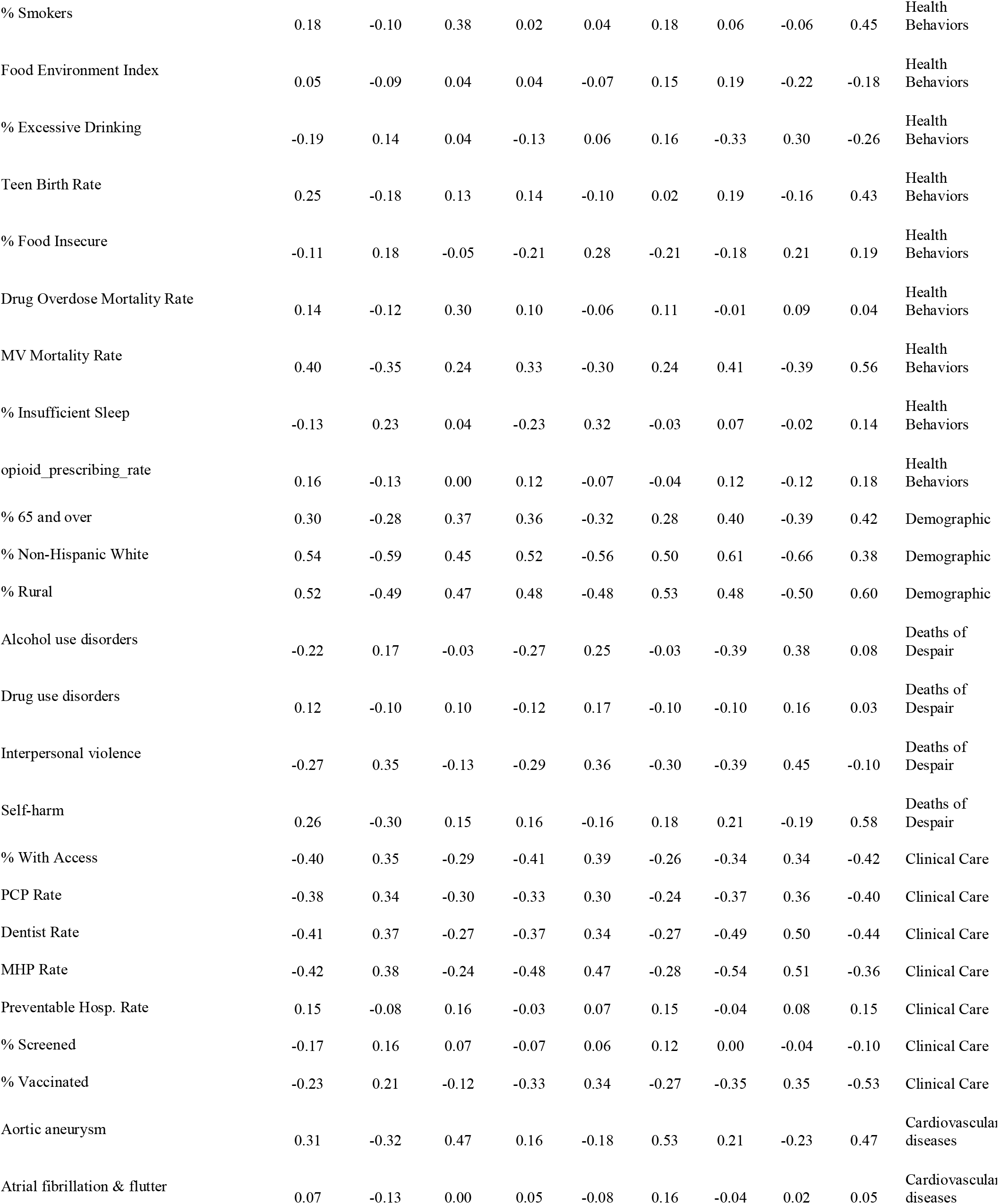
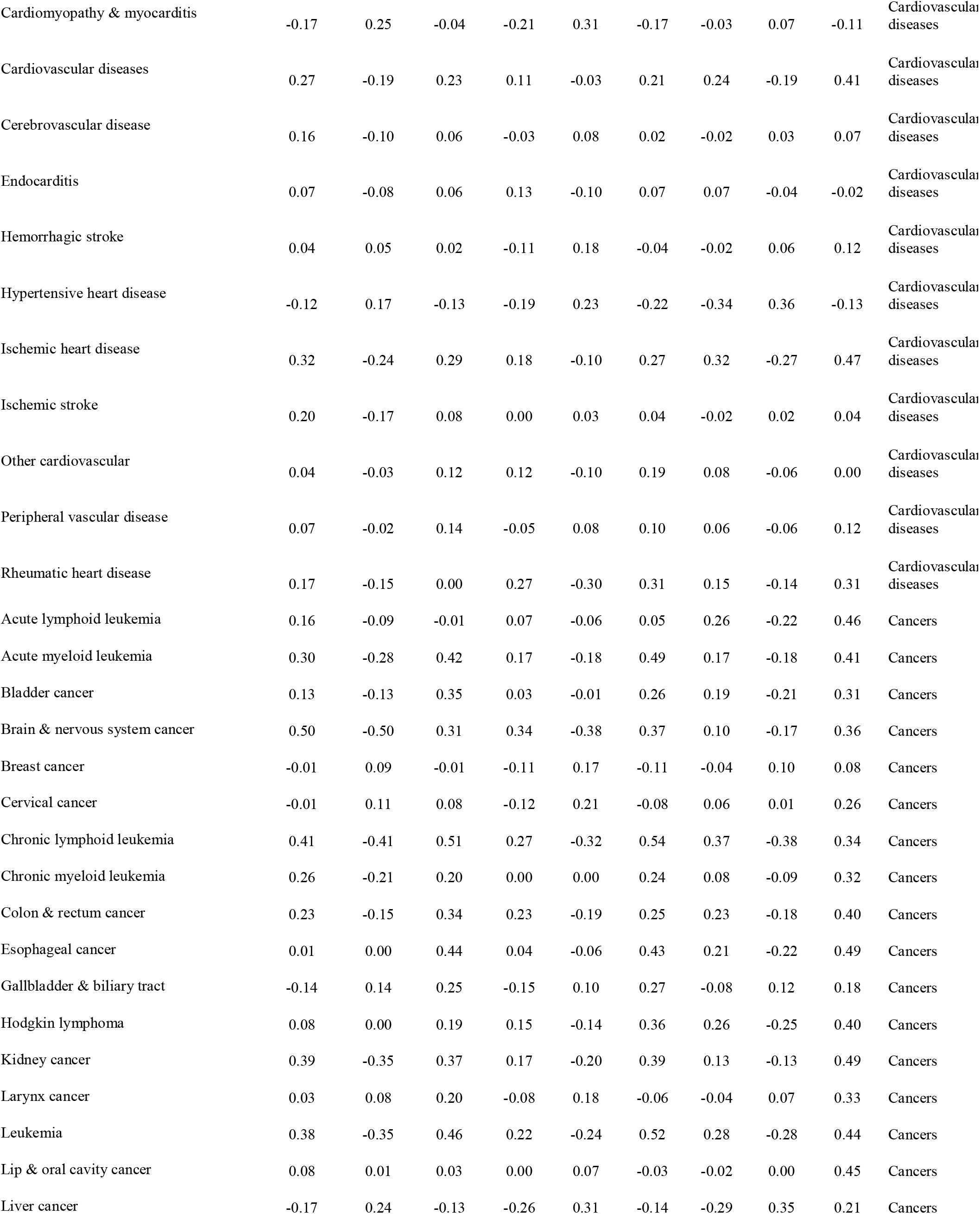
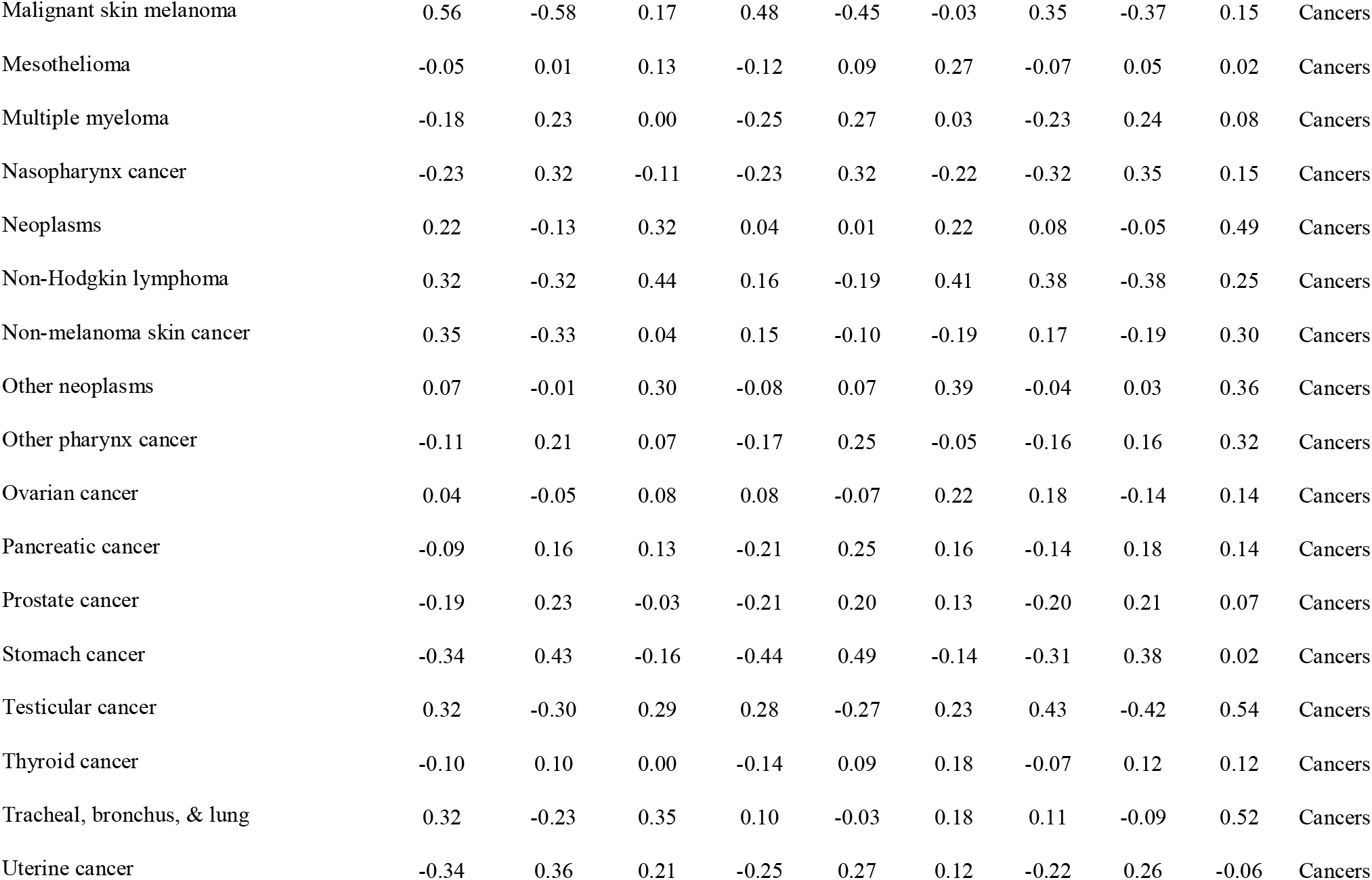
Quantile and mean comparisons of Republican and Democrat counties across public- health measures. Every county was assigned as either Republican or Democrat depending on the majority vote in 2016, and the mean, median, 1st quartile, and 3rd quartile values for different public health-related variables were calculated. The differences in these values for Republican and Democrat counties are presented in Table 2, along with the Student t-test statistics and p values for the mean comparisons.

Political voting data at the county level for presidential voting in 2012 and 2016 was collected from the MIT Election project.^23^ The margin shift was calculated by taking the difference in the Republican margin (Republican percentage of total vote minus Democrat percentage of total vote) from 2012 to 2016. We define Republican or Democratic counties as those that voted in favor of the Republican or Democratic candidate in 2016. Whenever possible, data from years as close to 2016 as possible was used (while 2016 data is available for most sources, the GBD data is from 2014).

Pearson correlations and confidence intervals for the correlations between all of the public health-related variables collected, the percentage of voters in the county that voted for Donald Trump or Hillary Clinton, and the Republican margin shift were computed from 2012 to 2016 (Table 1). Correlations for counties from all states, counties from battleground states (defined as states that could be reasonably won by either party), and counties from states that “flipped” from Democrat in 2012 to Republican in 2016, are presented. The battleground states are: Nevada, Colorado, Virginia, Florida, Michigan, Minnesota, Wisconsin, Iowa, New Hampshire, Ohio, and Pennsylvania. The flipped states are: Michigan, Pennsylvania, Wisconsin, Maine. In order to make Supplementary Table 1, we use the same data and structure as Table 1, except the correlations are instead weighed correlations, where the weights are equal to the base 10 logarithm of the county’s population.

Every county was assigned as either Republican or Democrat depending on the majority vote in 2016, and the mean, median, 1st quartile, and 3rd quartile values for different public health-related variables were calculated. The differences in these values for Republican and Democrat counties are presented in Table 2, along with the Student t-test statistics and p values for the mean comparisons.

The distributions of different public health variables for Republican and Democratic counties are presented in Fig 1. The dynamics of different public health variables over time for counties based on their 2016 political party are compared across the aggregated data (Fig 2). The Republican margin shift was then compared with different public health variables for counties in states that flipped from Democrat in 2012 to Republican in 2016, indicating the 2016 total number of votes and the 2016 election outcome for counties by the size and color of their points, respectively (Fig 3).

**Fig 1:**
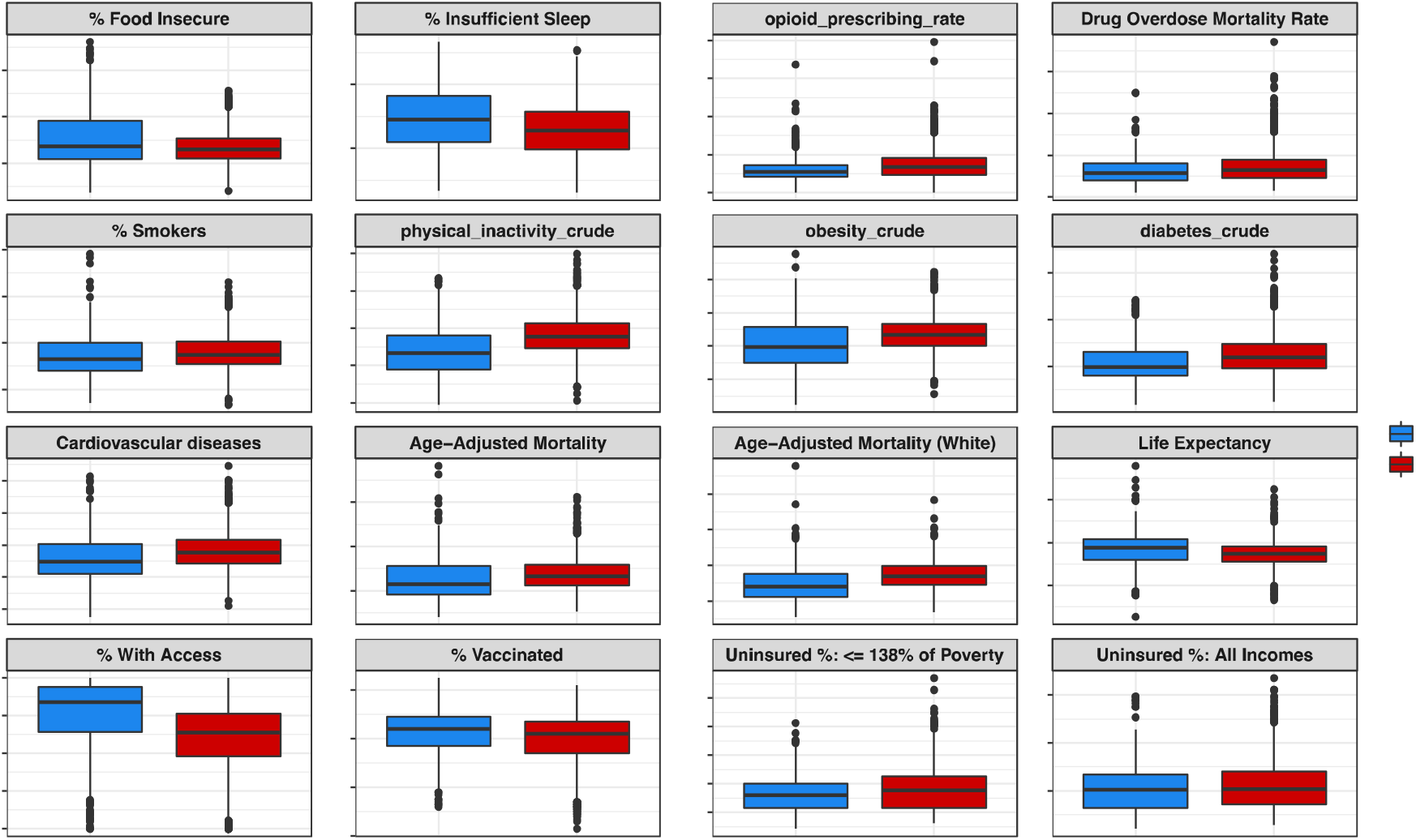
Boxplots comparing select public health variables for Democratic and Republican counties. As shown, there are higher rates of lifestyle factors like smoking, obesity, and physical inactivity, and chronic diseases that are affected by lifestyle like cardiovascular diseases and diabetes in Republican counties than in Democratic counties. Democratic counties also have higher life expectancy, insurance rates, and lower mortality rates than Republican counties. The percent of individuals who are food insecure or get insufficient sleep in Democratic counties is higher than in Republican counties.

## Results

Our analysis covers 3,156 counties from all 50 states and Washington DC, of which 2650 went Republican in 2016, and 506 went Democratic in 2016. These counties exhibit significantly different demographics: the average “% 65 and older” in Republican counties was 19.68% higher compared to Democratic counties; the average “% Non-Hispanic White” in Republican counties was 50.20% higher; the average “% Rural” in Republican counties was 93.91% higher; and the average “% with Some College” in Republican counties was 8.51% lower. These demographic differences are also driving healthcare differences. For example, Republican counties received 52% of Medicare funding (of which patients over 65 account for 85%^24^) in 2017, compared to 50.5% of spending in 2007. Additionally, the total number of non-elderly individuals with preexisting conditions in states that voted Republican in 2016 was 74.3 million, compared to 59.4 million in Democrat states. However, the average rate of non-elderly people with preexisting conditions in Republican states was 50% compared to 51% in Democratic states.

Table 1 shows Pearson correlations between different public health-related variables and the percentage of voters in counties who voted for Trump in 2016, for Clinton in 2016, and the Republican margin change from 2012 to 2016. These correlations were calculated for all states, for 2016 battleground states, and for states that flipped in 2016. For counties in all states, the percentage of votes for Trump had a correlation with the life expectancy of whites of -.42, and a correlation with physical inactivity of .36. Some of the variables that are highly correlated with the Republican margin shift in flipped states include mortality risk across all age groups, the percentage of people in the counties on Medicaid across all ages/sex groups, and the overall uninsured rate.

The median Republican county had 11% fewer residents who completed some college and the bottom and top quartiles were 6% and 11% less respectively. Table 2 shows percent differences between the 1^st^ quartile, median, 3^rd^ quartile and means for different health-related variables as well as t-statistics and p-values. The median Republican county had a 17% higher “injury death rate” and a 26% higher “% Disconnected Youth” rate. The median Republican county had a 50% higher rate of coal workers’ pneumoconiosis, 25% higher rate of chronic respiratory diseases, and a 32% higher rate of chronic obstructive pulmonary disease. The mortality risk was higher in every age group for the median, and bottom quartile counties, and lower for the age groups less than 65. The healthcare costs per capita were higher in Democratic counties (including Imaging Costs per Capita, Procedures Per Capita Actual Costs, Tests Per Capita Actual Costs, and Actual Costs per capita). There were consistently higher Medicaid participation rates among Democratic counties. The median uninsured % and uninsured % among individuals with less than 138% of the poverty line were higher in Republican counties by 2% and 11%, respectively. The insurance rates were much higher in Democratic counties among those with and without a high school education. The median Republican county had 2% lower life expectancy overall and 3% lower life expectancy among whites. Cancer rates were higher in most Republican counties than Democratic counties, with the exceptions of prostate, liver, and stomach cancers.

The median Republican county has a 13% higher obesity rate, a 21% higher diabetes rate, 19% higher physical inactivity rate, a 24% higher opioid prescribing rate, and a 6% higher smoking rate. Republican counties are older, with the median Republican county having 21% more individuals in the % 65 and over demographic. They are also whiter, with the median Republican county having a 69% greater rate in the % Non-Hispanic White demographic. Republican counties are more rural (median % rural rate is 234% higher for Republican counties), and access to care decreases in these counties accordingly: the primary care physician rate (ratio of population to primary care physicians) is 37% lower in the median Republican counties. Some of these health behavior, life expectancy, and health insurance rate differences presented in Table 2 are visualized in Figure 1.

Figure 2 shows the dynamics of healthcare and mortality in Democratic and Republican counties over time, visualizing the rates of different diseases, and mortality over time. Over the past 10+ years, life expectancy has changed at different paces, and has improved faster in Democratic counties. For health behaviors and chronic disease, since 2008, physical inactivity, diabetes, and obesity have become notably worse in Republican counties. While the mortality risk across all age groups has decreased overall since 1980, the mortality risk is now higher in the median Republican county compared to the median Democrat county for all age groups. Supplemental Figure 1 shows the clear growing differences between several health and life expectancy measures in the median Republican and Democrat county over time.

**Fig 2:**
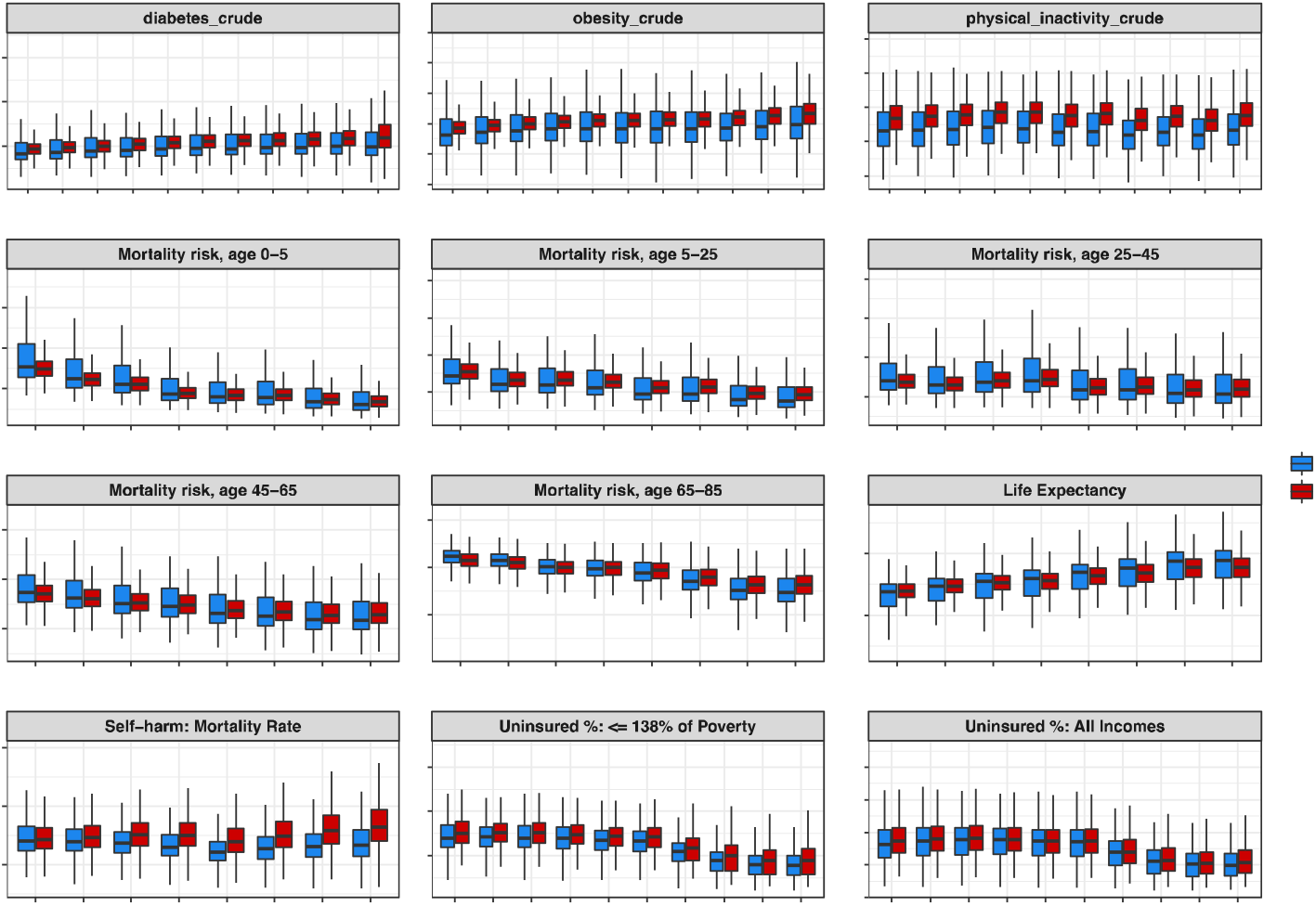
Boxplots comparing Democratic and Republican counties (defined by 2016 presidential election voting) over time for a number of public health variables. For diabetes, obesity, and physical inactivity, there has been a growing divide between Republican and Democratic counties between 2006 and 2016. For mortality risk across every age group, Democrat counties have improved more than Republican counties over the time period from 1980 to 2014.

Much of the 2016 election media narrative was focused on the rural, white, over 65 voters who supported Trump. Figure 3 shows health and demographic variables that are strongly correlated with the Republican margin shift in counties in the states that flipped from Democrat to Republican in 2016. We can also see that obesity, diabetes, physical inactivity and smoking are all highly correlated with the Republican margin shift. There is a strong negative correlation between the life expectancy of whites and the Republican margin shift in these flip states.

**Fig 3:**
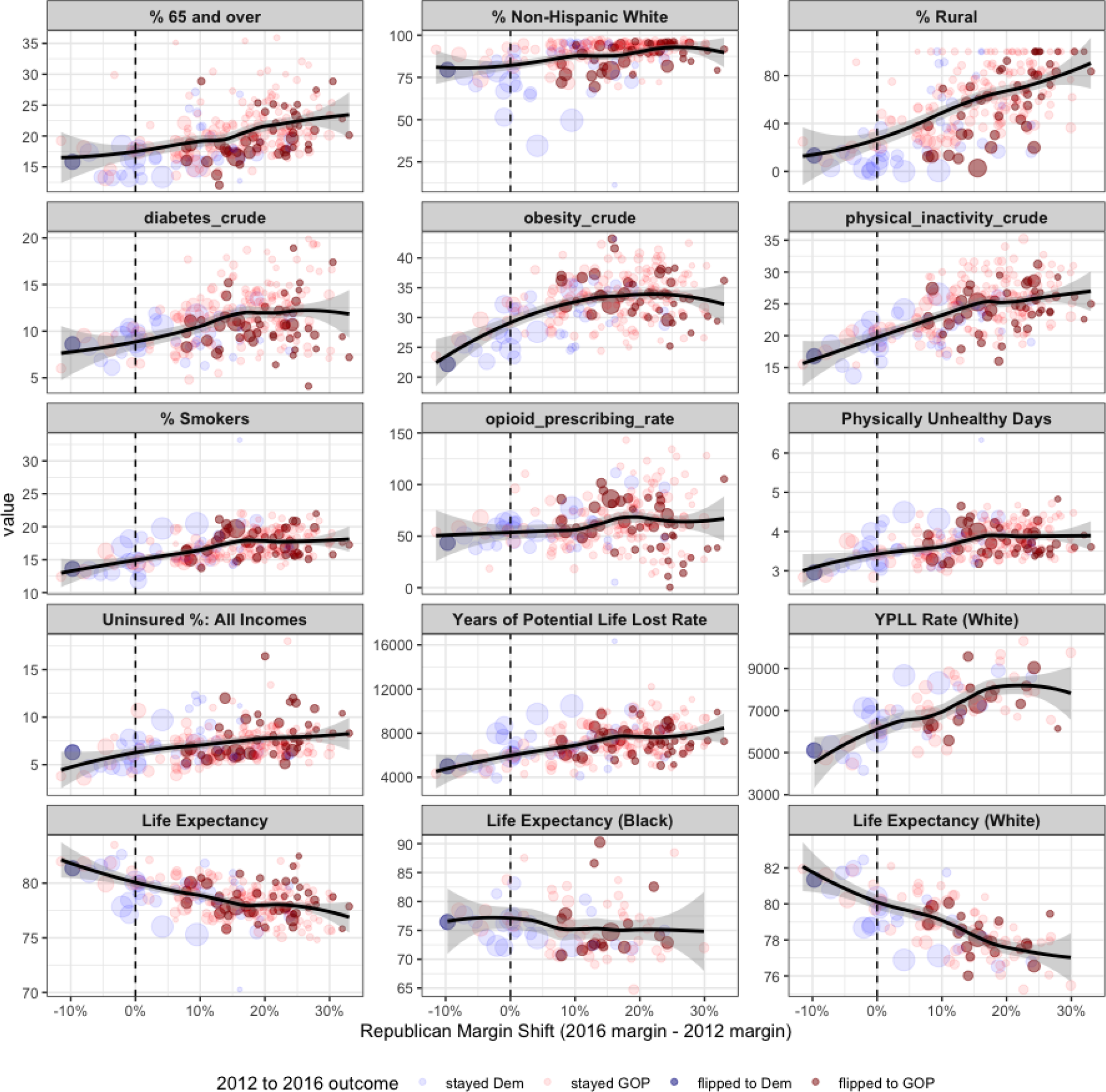
Scatterplots for all counties in the 4 states that flipped from Democrat to Republican in 2016 (Michigan, Pennsylvania, Wisconsin, Maine), showing the Republican margin shift on the x axis, and different demographic and public health related variables on the y axis. Counties are sized by the total number of votes made in the 2016 election, and they are colored by the 2012 and 2016 outcomes. The top row includes variables frequently discussed in the narrative around the electoral shift in these states, including the percentage of Non-Hispanic Whites in the county. There is a clear relationship between the obesity rate, physical inactivity rate, smoking rate, and life expectancy and the Republican margin shift in these states.

Most of the states that participated in Medicaid expansion (though not all) voted Democrat in 2016. States that expanded Medicaid improved the insurance rates of their states and tended to have higher insured rate changes than states that did not, although there are a few exceptions (such as Florida and Idaho) that experienced large changes over this period without expansion. Medicaid expansion was particularly impactful for the insurance rates of individuals making less than 138% of the poverty line. Supplementary Fig 2 shows insurance rate changes from 2008 to 2017 (capturing the impact of the ACA) for counties in States that did and did not implement Medicaid expansion. Each point represents a county. For this group, Medicaid expansion directly improved the insurance rates of states.

## Discussion

In this retrospective cohort study, we found statistically significant relationships between a number of health measures and the political voting patterns of counties in 2016 and over the last three decades. By calculating the median difference between counties that voted Democrat or Republican in the 2016 election, we found that residents of counties that voted Republican in the last presidential election have increased median incidence cardiovascular disease (11% median difference), diabetes (21%), obesity (13%), self-harm (22%), decreased median life expectancy (2%), and physical activity (19%) compared to residents of counties that voted Democrat. Collectively, these data indicate that counties that voted Republican in the 2016 election are “sicker” than those that voted Democrat.

The trends in Fig 2 show increases over time in counties that voted Republican in 2016 compared to Democrat in negative health outcomes such as diabetes and obesity concomitantly with decreases in life expectancy. This indicates that these counties have experienced an overall worsening in quality of health over time. It is important to note that these are not necessarily counties that have voted Republican in previous elections. The Affordable Care Act (ACA) was the signature legislative accomplishment of the Obama administration. Yet during his presidency, counties that ultimately voted Republican in the 2016 election continued to experience an overall worsening of healthcare measures, despite improved insurance rates. This worsening in the context of the ACA may be one of a number of factors contributing to individual voting decisions for Trump in 2016.^25^

As the incidence of these chronic diseases grows, so does the overall cost of treating them. 90% of the nation s annual healthcare expenditures of $3.5 trillion is for people with chronic and/or psychiatric conditions. Diabetes accounts for $237 billion every year, with 30 million Americans affected and 84 million with prediabetes; obesity directly costs $147 billion a year; lack of physical activity costs $117 billion annually; smoking leads to $170 billion in direct medical costs; and heart disease and stroke cost $199 billion per year and lead to $131 billion in lost productivity.^26^ As patients live with these chronic diseases for longer, the number of complications for which treatment will be necessary will increase, further increasing costs. As these costs rise, it is likely that attempts will be made to curb healthcare spending. This will trap patients in these vulnerable counties in a vicious cycle: lack of access will lead to increased incidence of chronic disease, which will increase healthcare costs per patient, which will further increase total healthcare expenditure. In light of proposed measures amounting to healthcare austerity, as well as the evidence of the deleterious effects of austerity on health in general, more targeted and effective healthcare spending (rather than cuts) is warranted.^27^

The majority of Medicare funds (52%) go to counties that voted Republican in 2016. Nevertheless, the Trump administration is mulling large cuts to Medicare and other entitlement programs.^28^ Additionally, the majority of individuals with preexisting conditions live in states that voted Republican in 2016 (55.5%), running counter to the Trump administration’s policies that weaken laws protecting those with pre-existing conditions.^29^

Our analysis showed trends between voting patterns and health outcomes, but it is also important to study patterns between health policy and political party affiliation. One direct consequence of political party affiliation on health policy is Medicaid expansion, which many more Democratic states adopted. All together, our data indicate that the health needs of certain areas are not being adequately addressed by their elected officials. States that expanded Medicaid were associated with a number of positive health measures, including improved access to care, better glucose monitoring in diabetes, better hypertension control, reductions in rates of major ^30,31,32^ postoperative morbidity, and reductions in preventable hospitalizations. The 14 states that have not expanded Medicaid (Alabama, Florida, Georgia, Kansas, Mississippi, Missouri, North Carolina, Oklahoma, South Carolina, South Dakota, Tennessee, Texas, Wisconsin, Wyoming) have overall lower median insured rates among those making 138% below the federal poverty level compared to states that expanded Medicaid. Lawmakers in states that have not expanded Medicaid have claimed doing so would harm the state economy.^33,34^ It has been demonstrated that Medicaid expansion has saved at least 19,200 lives and, conversely, 15,600 deaths have occurred as a result of non-expansion.^35^ In light of these data, as well as those indicating states that had Medicaid expansion showed improved health measures, it is clear that states that did not expand Medicaid harm their constituency by failing to do so. The impact of health has been previously shown to affect political participation, with people in poor health being less likely to vote.^36^ Researchers have also looked at the public health voting patterns in the US senate and found that Democratic senators averaged 59.1% points higher in voting accordance with public health policy recommendations compared to Republican Senators, suggesting that Republican senators diverge significantly from the recommendations of the American Public Health Association (APHA).^37^ Given the large the differences in health behaviors of Democrat and Republican counties with respect to physical inactivity, obesity, and smoking, the policy differences should be fully examined. It is worth systematically exploring the extent to which political party affiliation affects public health policymaking and the impact this has on public health.

As of late May 2020, the COVID-19 pandemic has affected Democratic and Republican counties differently. The most cases and deaths from the virus have occurred in dense urban areas, which lean Democrat, including the epicenter in New York City. The New York Times estimated that counties Trump won in 2016 reported 27 percent of the cases and 21 percent of the deaths.^38^ This divide is further highlighted by the partisan nature of policies implemented by elected officials (i.e. governors) in response to the crisis.

Healthcare is arguably the central topic of the 2020 election, with COVID-19 serving as an avatar for the politicization of healthcare. Medical care during the pandemic, risk of complications during the pandemic, perceptions of the pandemic, behavior change during the pandemic, the government response during a pandemic, and the social safety net during the pandemic, have all been divided along partisan lines.

This study has several limitations. The first is that we cannot attribute voting behavior to individual Democrat and Republican voters. While we have examined the voting patterns at the county level, it is true that voter turnout in the United States is low (55.7% in the 2016 election) compared to other developed countries and varies between demographics, so whether the results in a given county reflect the true preferences of its residents is questionable. Similarly, the majority of lower socioeconomic individuals do not vote, which skews the data towards those who can, who may also be healthier on average. These data do not indicate the healthcare differences between individual voters and should not be understood as a reflection on individual party members.

Here we find strong relationships between voter party affiliation and health outcomes. These outcomes stem from both individual mechanisms (like the self-expressed lower priority of health issues in republican voters) as well as institutional aggregate measures (e.g. ACA and Medicare expansion choices falling along party lines). Although polarization and partisanship are increasing in western democracies, the link between partisanship and health is particularly strong in the US, and our work suggests that it is in the public interest to further study the mechanisms that link partisanship to health outcomes in an attempt to decouple political affiliation and health in the future.

## Data Availability

All of the data analyzed in this manuscript is available at the following link: https://zenodo.org/record/3936108#.Xyc5O_hKh_Q with the DOI number 10.5281/zenodo.3936108.

## Data and code availabillity

The analysis and code from this manuscript can be found at the following link: https://github.com/tymor22/Health-and-Politics/. All of the data analyzed in this manuscript is available at the following link: https://zenodo.org/record/3936108#.Xyc5O_hKh_Q with the DOI number 10.5281/zenodo.3936108.

## Acknowledgments

We thank Kevin Aslett for a careful reading of the manuscript.

---

**Supplemental Fig 1.**
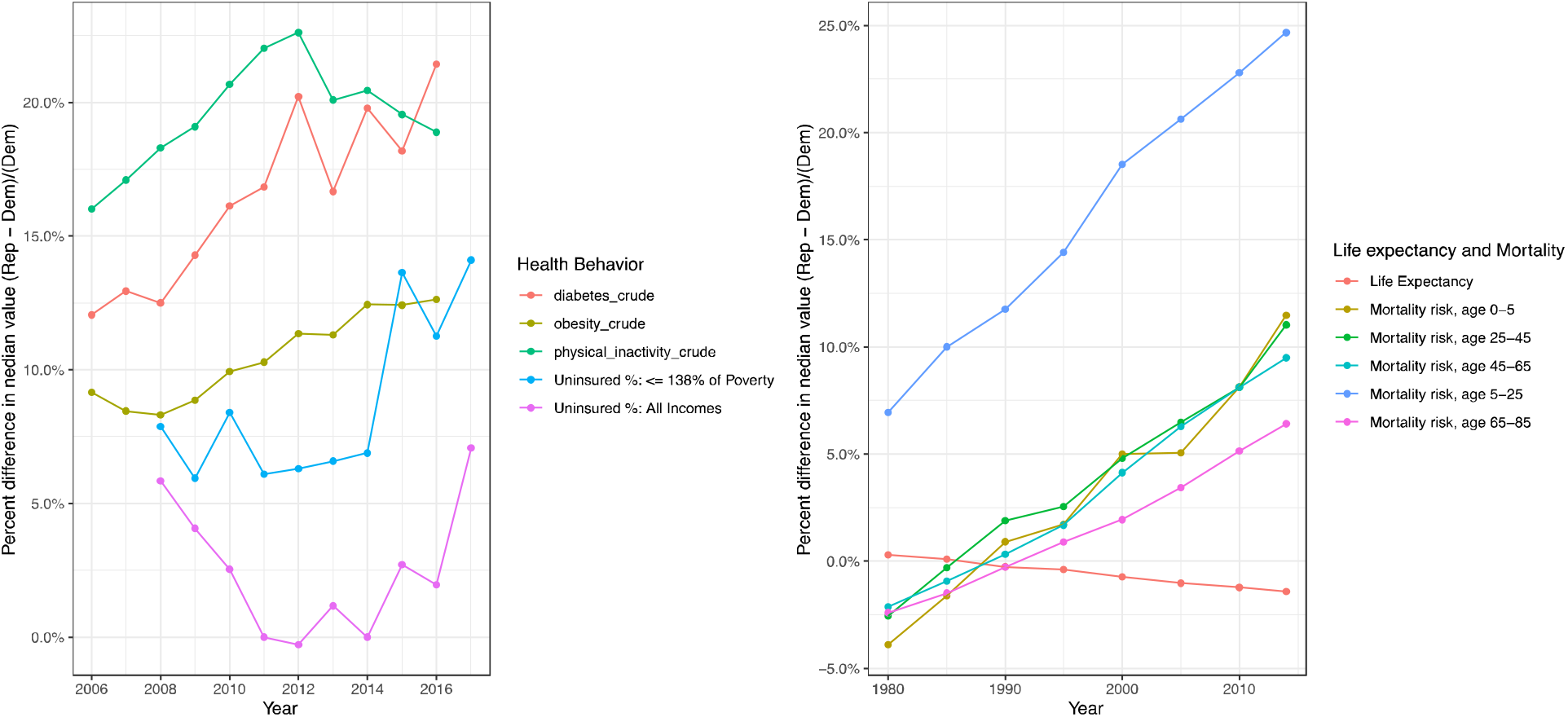
Percent differences in the median value of Republican and Democratic counties for select life expectancy, mortality, and health behavior measures. The median Republican county has experienced sustained increases in mortality risk across every age group compared to the median Democratic county between 1980 and 2014; this manifests in worse life expectancy for the median Republican county over time. Diabetes, obesity, physical inactivity, and uninsurance rates in the median Republican county are higher than in the median Democratic county between 2006 and 2017, and this difference is growing.

**Supplemental Fig 2.**
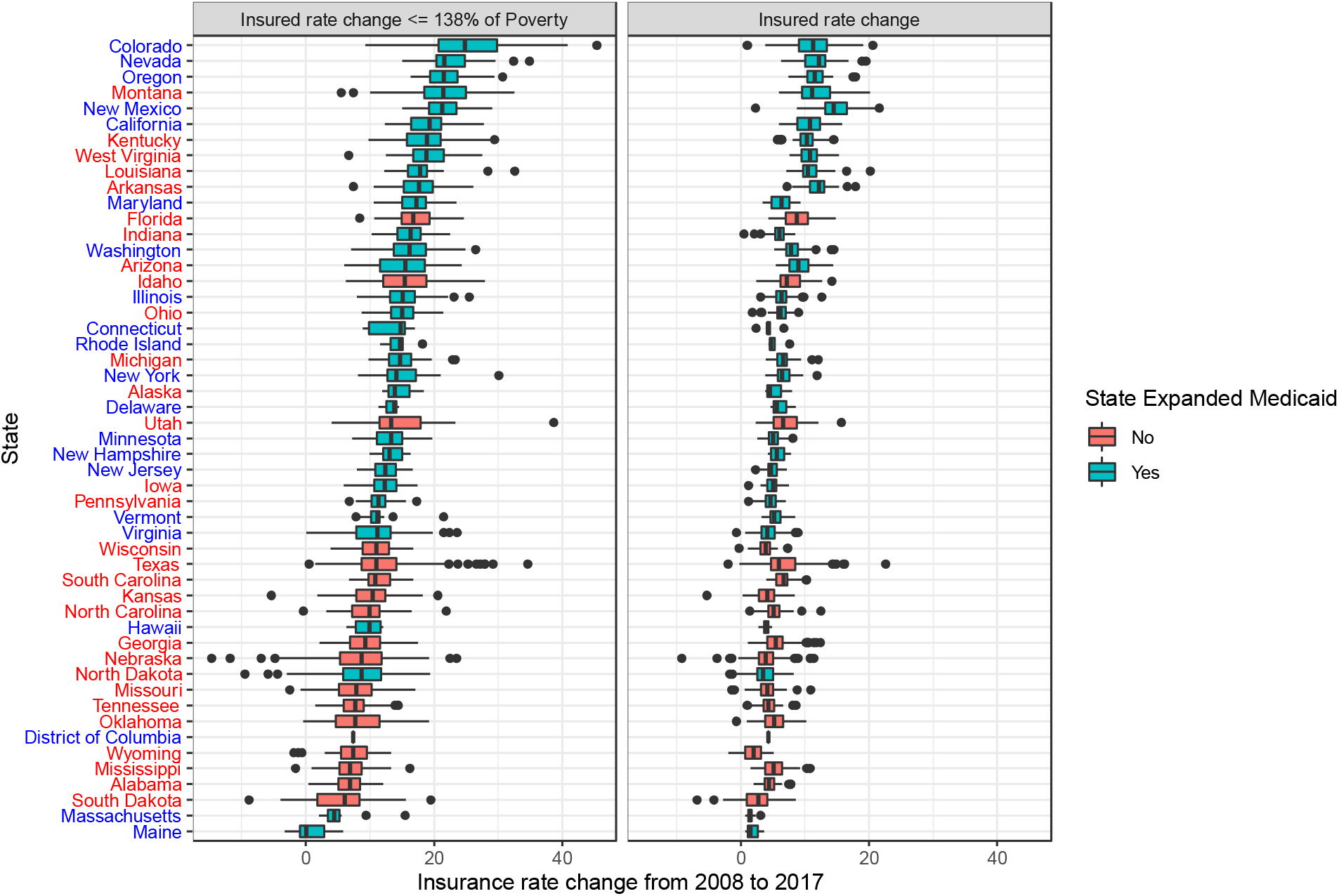
Boxplots of insurance rate changes between 2008 and 2017 for counties in states. Boxplots are filled by whether the state expanded Medicaid, and state names are colored by the 2016 political party. States that expanded Medicaid experienced higher insurance rate changes during this time period, indicating the positive impact of the policy.

**Supplemental Table 1.**
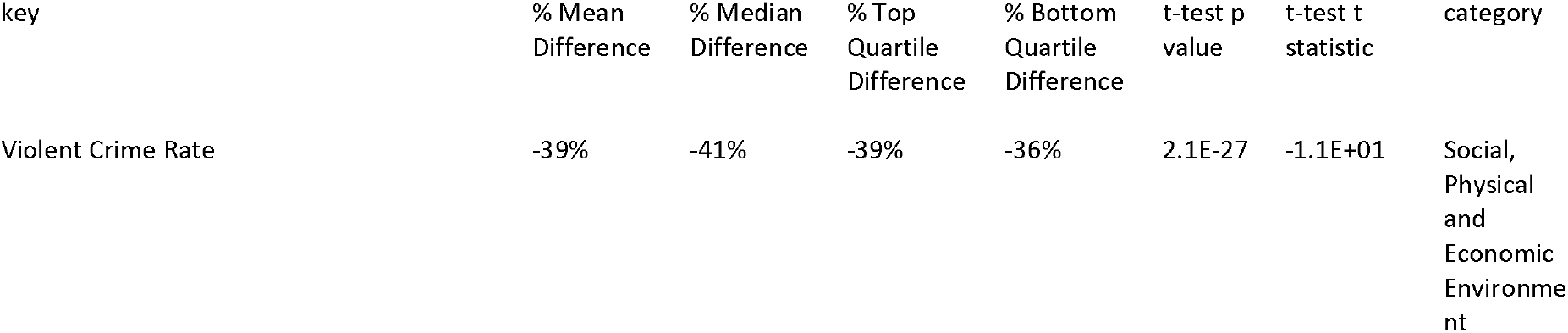
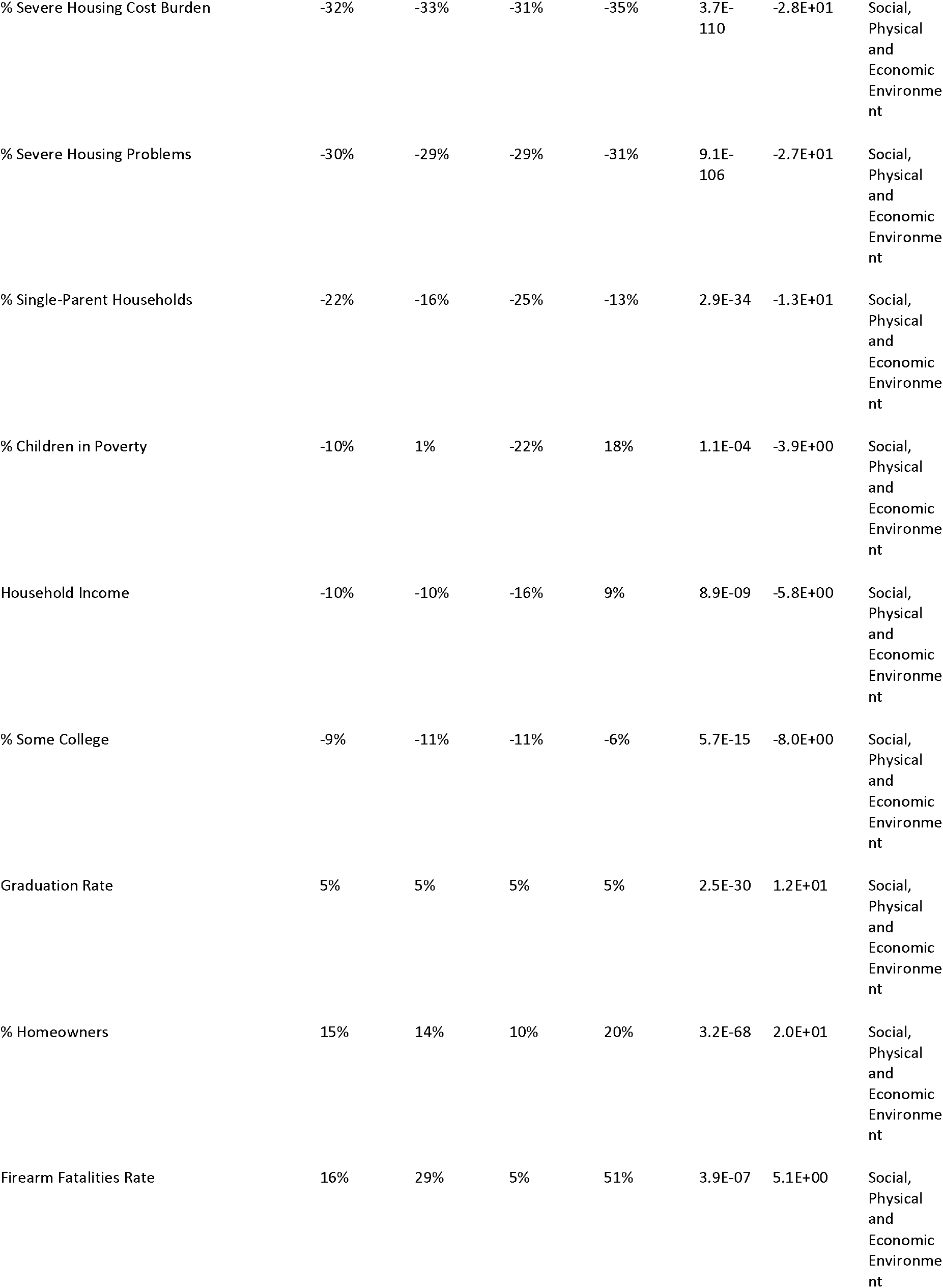
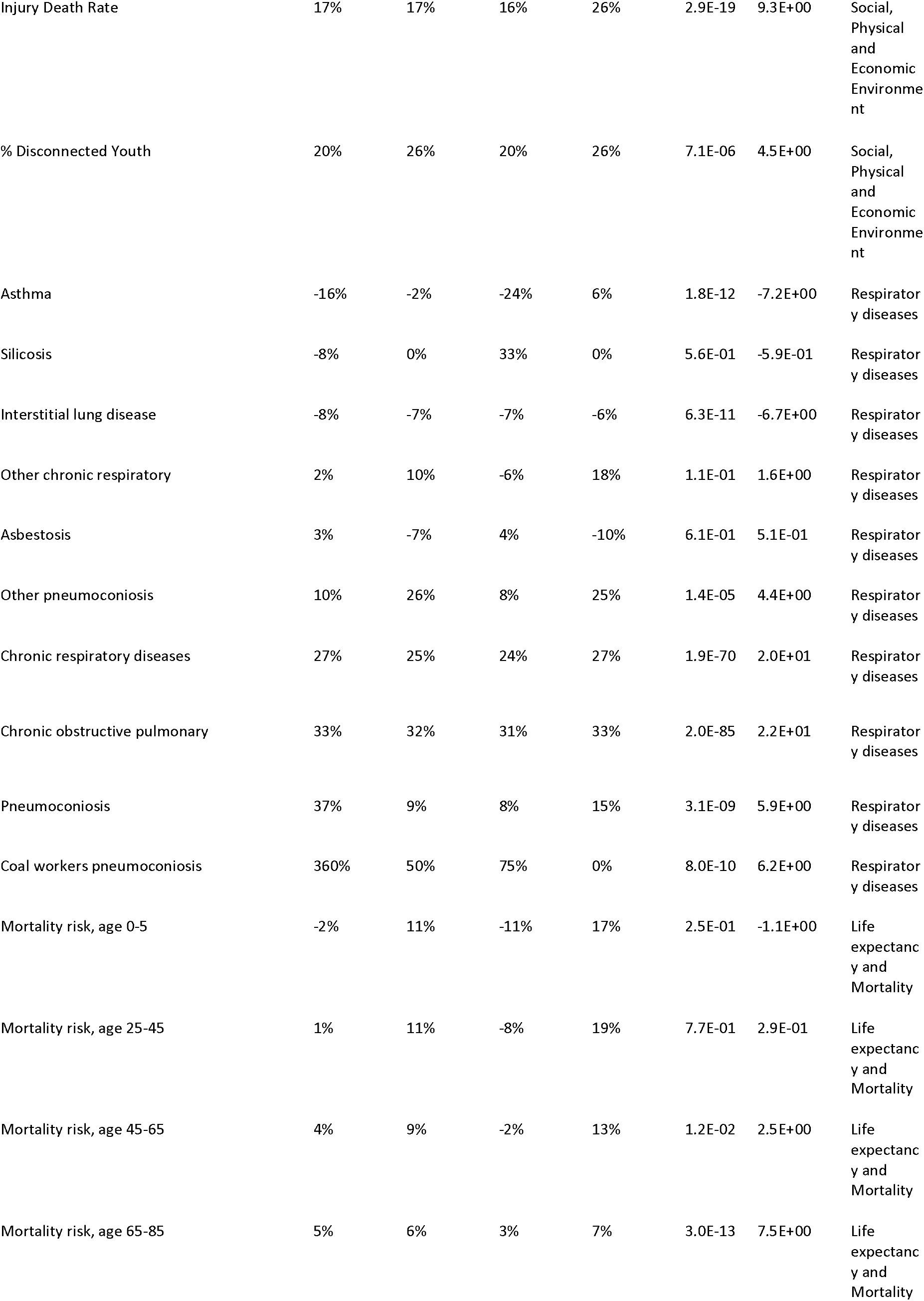
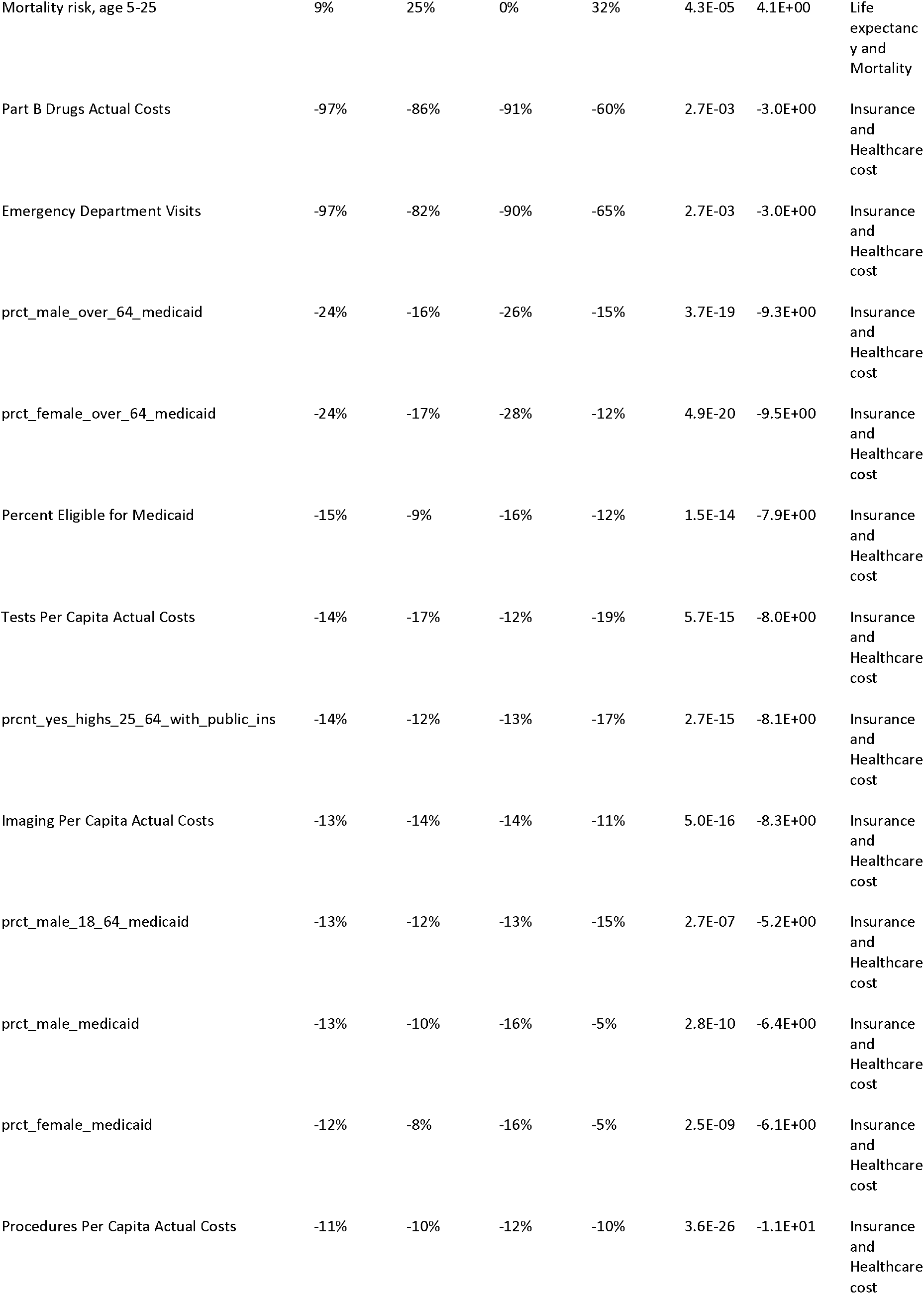
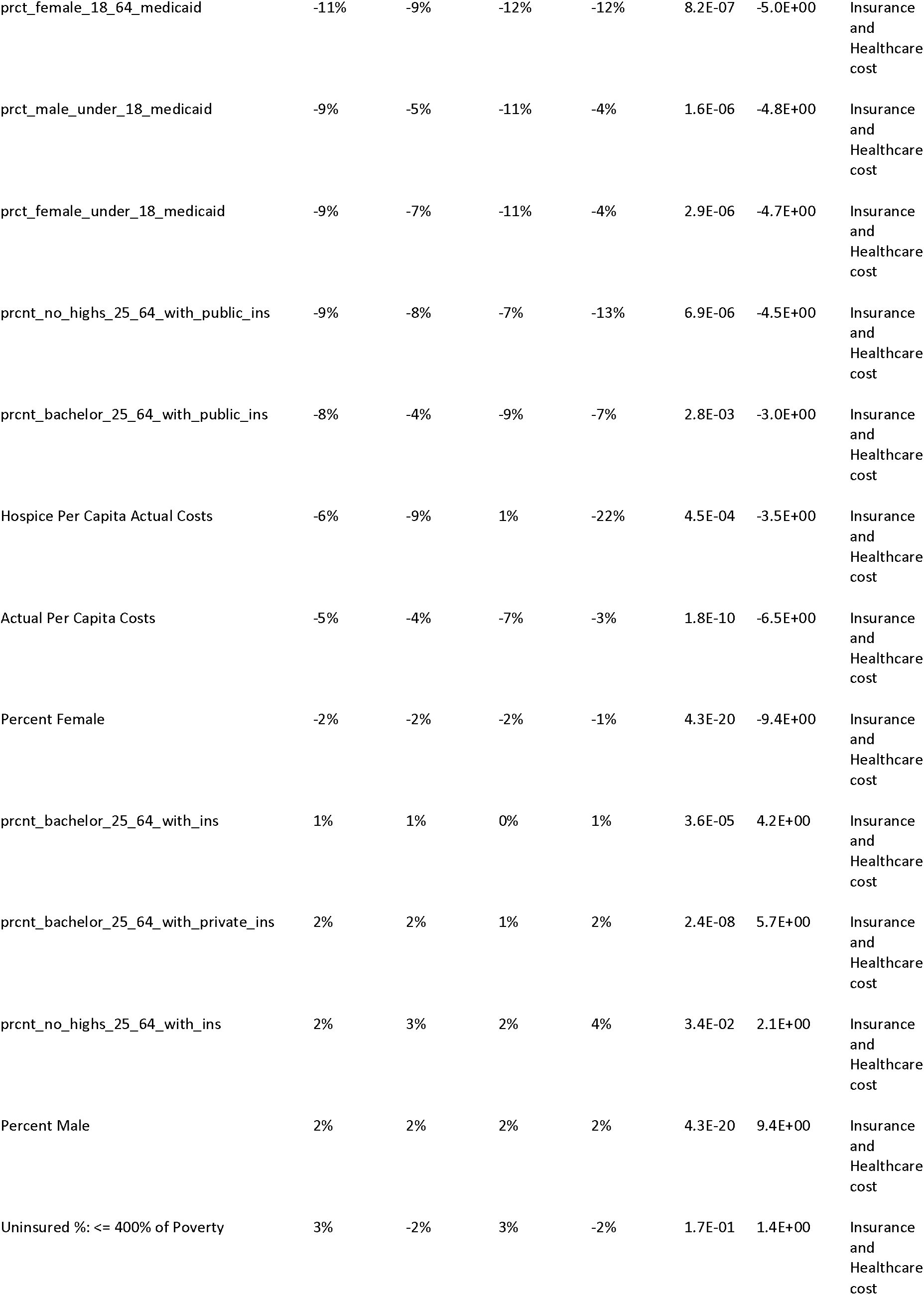
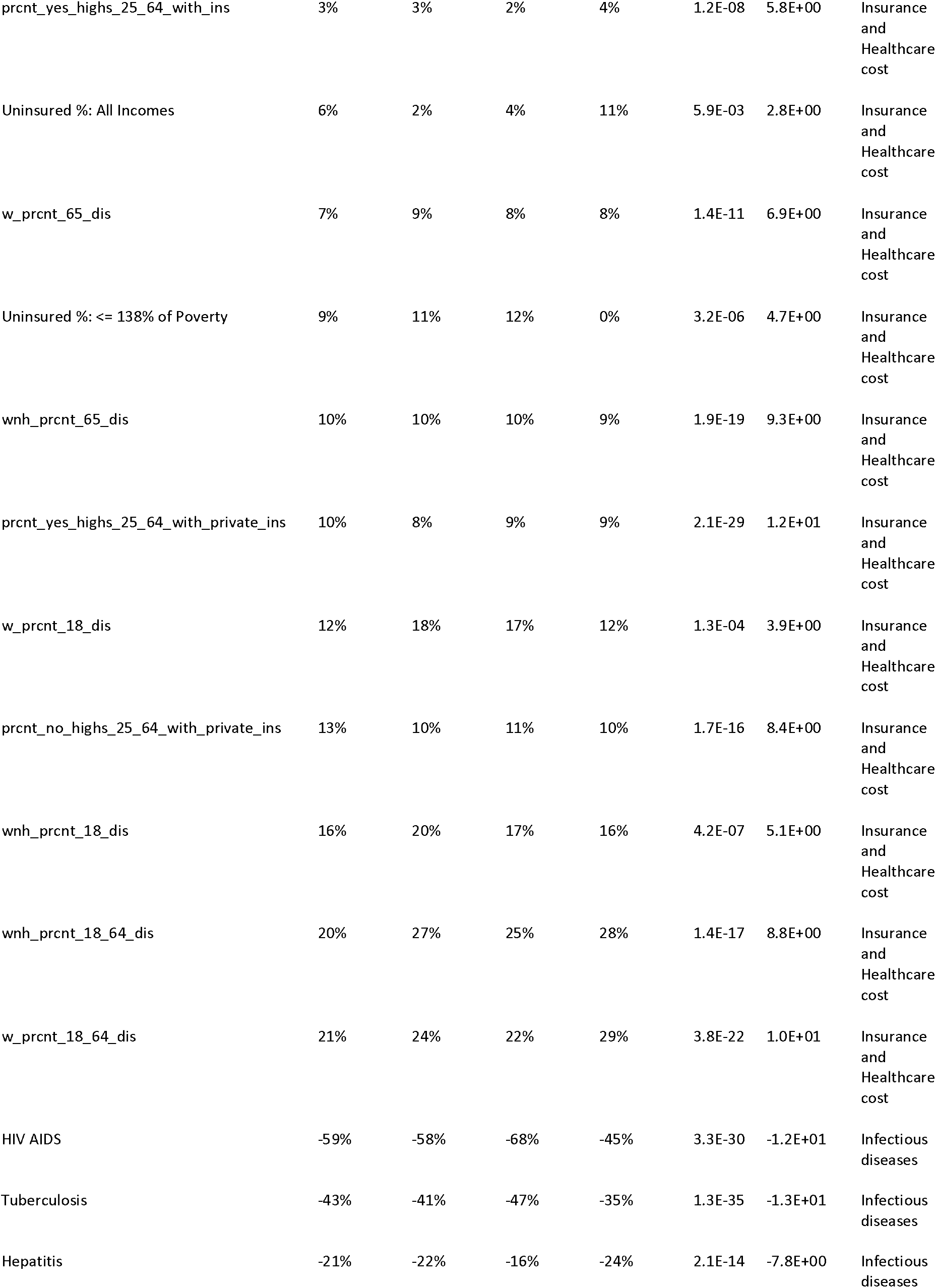
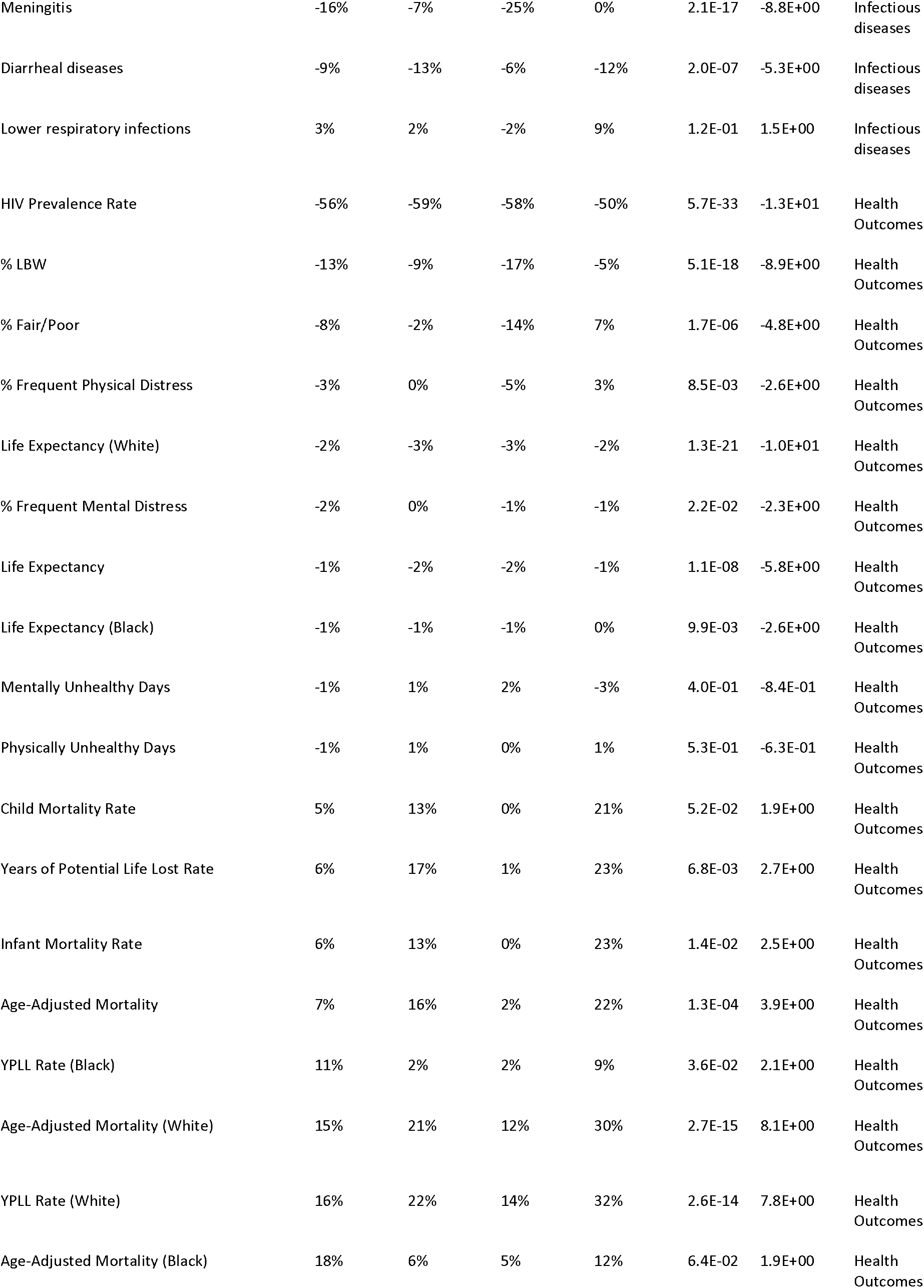
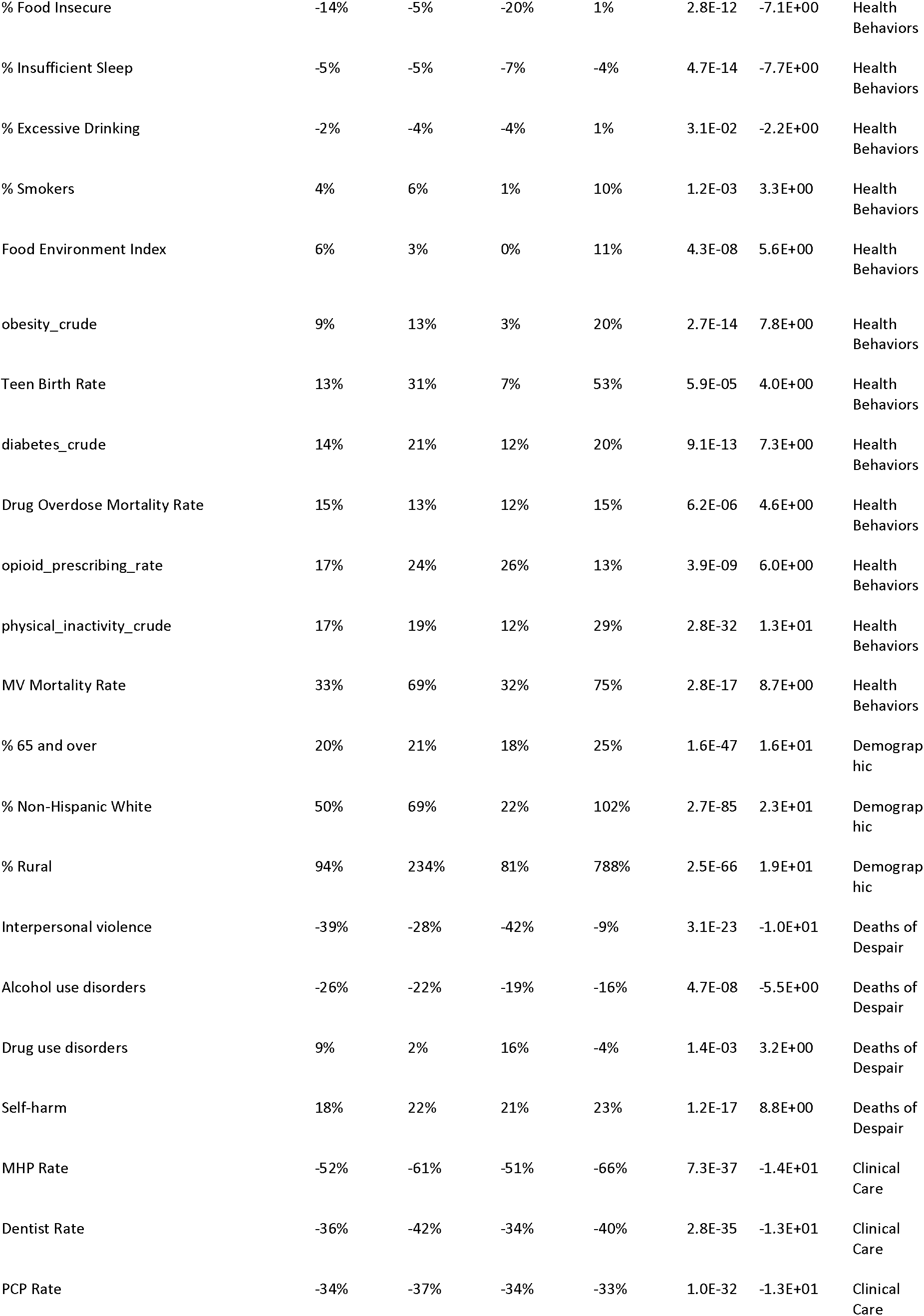
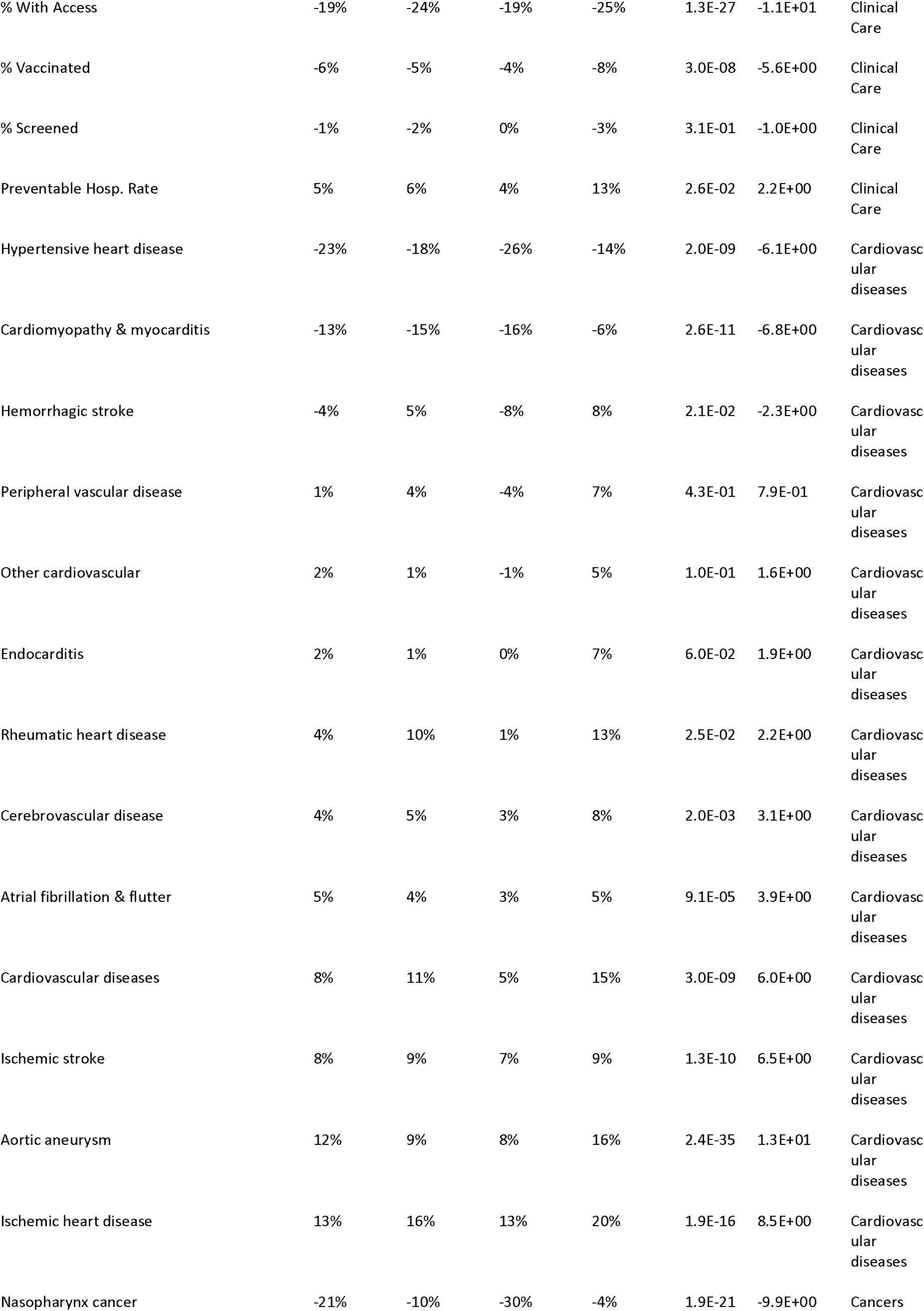
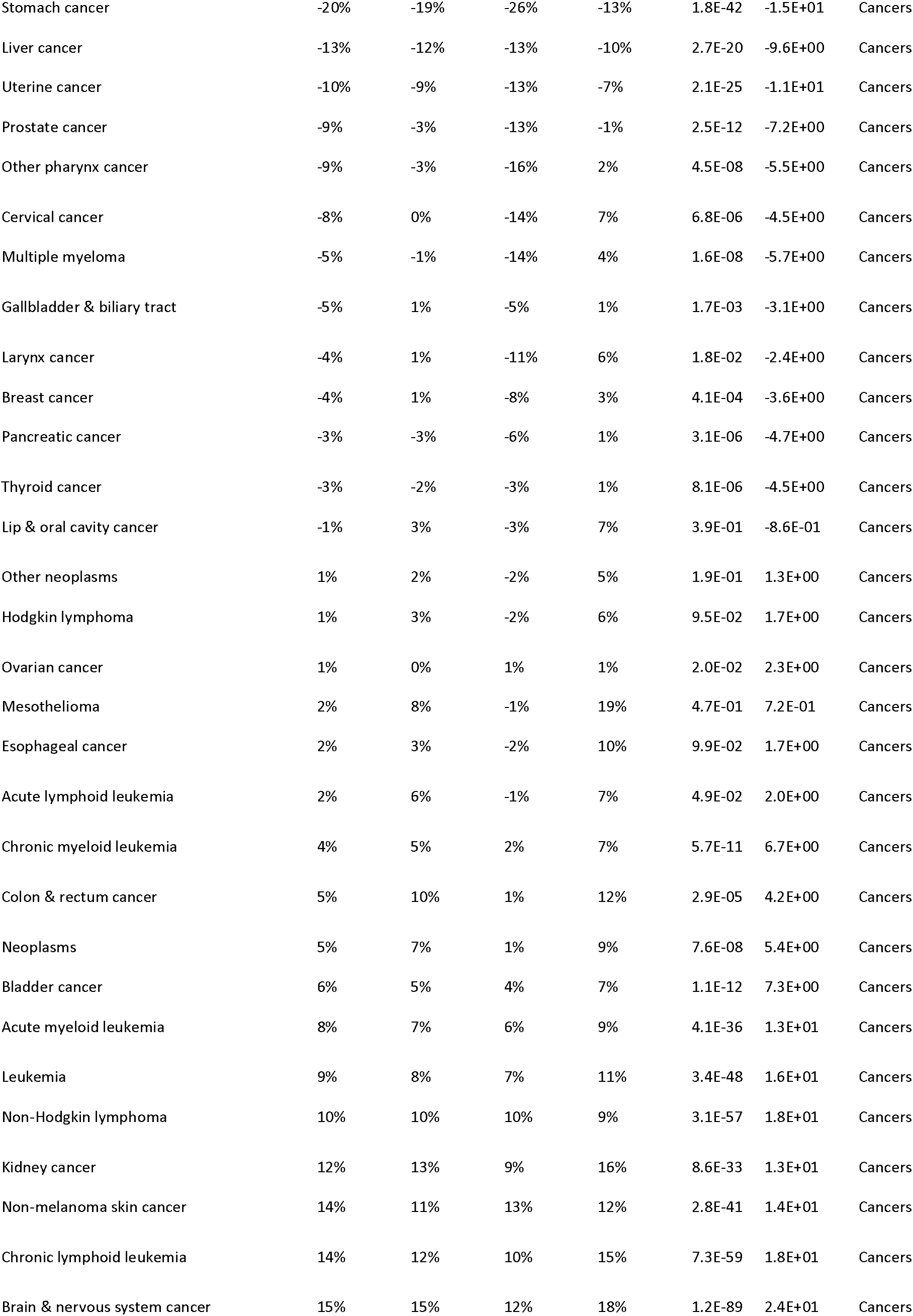
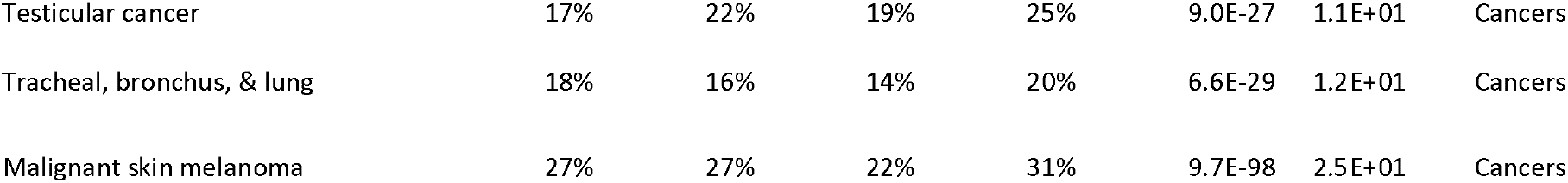
Weighted Pearson correlations (weighted by the log 10 of the county population) between different public health-related variables with the percentage of voters in thecounty that voted for Donald Trump or Hilary Clinton, and the Republican margin shift (from 2012 to 2016).

